# Clinical Impact of Panel Based Error Corrected Next Generation Sequencing versus Flow Cytometry to Detect Measurable Residual Disease (MRD) in Acute Myeloid Leukemia (AML)

**DOI:** 10.1101/2020.08.23.20180372

**Authors:** Nikhil Patkar, Chinmayee Kakirde, Anam Fatima Shaikh, Rakhi Salve, Prasanna Bhanshe, Gaurav Chatterjee, Sweta Rajpal, Swapnali Joshi, Shruti Chaudhary, Rohan Kodgule, Sitaram Ghoghale, Nilesh Deshpande, Dhanalaxmi Shetty, Syed Hasan Khizer, Hasmukh Jain, Bhausaheb Bagal, Hari Menon, Navin Khattry, Manju Sengar, Prashant Tembhare, Papagudi Subramanian, Sumeet Gujral

**Affiliations:** Haematopathology Laboratory, ACTREC, Tata Memorial Centre, Navi Mumbai, India; Homi Bhabha National Institute (HBNI), Mumbai, India; Dept of Cytogenetics, ACTREC, Tata Memorial Centre, Navi Mumbai, India; Adult Haematolymphoid Disease Management Group, Tata Memorial Centre, Mumbai, India; Haemato-Oncology, CyteCare Cancer Hospital, Bangalore, India

**Keywords:** Error Corrected Next Generation Sequencing, Measurable Residual Disease in AML, Unique Molecular Identifier based MRD Detection, Acute Myeloid Leukemia Risk of Relapse

## Abstract

We accrued 201 patients of adult AML treated with conventional therapy, in morphological remission and evaluated MRD using sensitive error corrected next generation sequencing (NGS-MRD) and multiparameter flow cytometry (FCM-MRD) at the end of induction (PI) and consolidation (PC). Nearly 71% of patients harbored PI NGS-MRD and 40.9% harbored PC NGS-MRD (median VAF 0.76%). Patients harboring NGS-MRD had a significantly higher cumulative incidence of relapse (p=0.003), inferior overall survival (p=0.001) and relapse free survival (p<0.001) as compared to NGS-MRD negative patients. NGS-MRD was predictive of inferior outcome in intermediate cytogenetic risk and demonstrated potential in favorable cytogenetic risk AML. Patients who cleared PI NGS-MRD had a significantly improved survival as compared to patients who became negative subsequently indicating that kinetics of NGS-MRD clearance was of paramount importance. NGS-MRD identified over 80% of cases identified by flow cytometry at PI time point whereas FCM identified 49.3% identified by NGS. Only a fraction of cases were truly missed by NGS as compared to FCM-MRD. NGS-MRD emerged as the most important independent prognostic factor predictive of inferior outcome (p<0.001). We demonstrate a widely applicable, scalable NGS-MRD approach that is clinically informative and advantageous when compared to FCM-MRD in AML treated with conventional therapies.

## Introduction

Acute Myeloid Leukemia is a disease characterized by heterogeneous biology resulting in varying clinical outcomes including relapse. ^1,2^ There are limited novel treatment options such as targeted therapies using *FLT3* or *IDH2* inhibitors and most patients are treated based on the presumptive risk of relapse. ^3,4^ This risk adapted therapy typically considers standard cytogenetic and molecular variables as recommended by the European LeukemiaNet. ^5^ Based on this risk, patients are recommended treatment with conventional (induction and consolidation) regimens or offered intensive therapy such as allogeneic bone marrow transplantation (BMT) after achievement of remission. Amongst these, the largest group remains as intermediate risk AML characterized by non-uniform clinical outcomes.

The monitoring of a patient’s response to chemotherapy, called, measurable residual disease (MRD) is one of the most important predictors of outcome in hematological malignancies. Several investigators have demonstrated the clinical utility of flow cytometry based MRD detection (FCM-MRD) in AML at early chemotherapy time points as well as in a pre-transplant setting.^6-14^ Although universally applicable, FCM-MRD suffers from suboptimal ability to predict relapse in AML compared to precursor B lineage acute lymphoblastic leukemia. A diverse array of sensitive molecular methods have been used to detect MRD in AML such as real time PCR^15^ and droplet digital PCR.^16^ These are useful for monitoring of individual gene mutations such as AML with mutated *NPM1*^17,18^ and chimeric gene fusions such as *RUNX1-RUNX1T1*.^19^ Next generation sequencing (NGS) is a promising tool for sensitive MRD monitoring and has been used successfully to monitor NPM1^20,21^, *RUNX1*^22^ and FLT3^23^ mutations as well as chimeric gene fusions.^24,25^ However, these methodologies can be utilized in only a subset of patients that harboured these mutations at diagnosis and this strategy discounts for clonal heterogeneity and evolution which are relevant to the pathogenesis of AML.

DNA based focussed target enrichment strategies (gene panels) are an attractive solution to detect MRD using NGS (NGS-MRD) in a mutation rich disease such as AML.^26,27-29^ However, short read sequencers are inherently prone to base calling errors limiting variant calling at 3-5% variant allele fraction (VAF).^30^ Although acceptable for diagnostic molecular pathology, this is undesirable assay performance for the detection of MRD. Error corrected sequencing involves the physical incorporation of random oligonucleotides or unique molecular identifiers (UMI) at the library preparation stage prior to amplification of DNA. This allows us to tag individual DNA molecules with an unique molecular fingerprint.^31,32^ Such approaches have been used for myelodysplastic syndromes^33^ and for pre transplant MRD monitoring of AML as demonstrated by Thol and colleagues.^34^ Thol utilized a sensitive patient specific mutation tracking approach using UMI based MRD detection. Although applicable to a broad spectrum of AML mutations, a tailored approach poses logistical and regulatory hurdles towards prospective MRD testing especially for early MRD timepoints.

In this study, we have evaluated the clinical utility of error corrected NGS to detect MRD in AML using single molecule molecular inversion probes (smMIPS).^31,35^ Each smMIP contains an 8 bp UMI and binds to a single molecule of DNA. Using consensus sequence-based variant calling we can detect somatic mutations including small indels in a sensitive manner. We demonstrate that error corrected NGS-MRD at early timepoints in therapy is significantly predictive of outcome in patients of AML treated with conventional therapies. Furthermore, we systematically compare multicolour FCM-MRD with error corrected NGS-MRD and assess the clinical utility of these two assays in a cohort of AML.

## Methods

1. PATIENTS
  a. *Patient Accrual and Initial work up:* The study was approved by the institutional ethics committee (IEC-III project 163) and participants were accrued after informed consent. A total of 393 adult patients of AML, diagnosed as per 2008 WHO criteria, were accrued in this study over a period of six years (Feb 2013 to May 2019). Cytogenetic (FISH and karyotyping) workup was as previously described.^9,21^ Somatic mutations at diagnosis were evaluated using a smMIPS based 50 gene myeloid panel as described previously by our group. ^36^ Of these, the smMIPS MRD panel (see below) was applicable to 83.2% patients [327 out of 393 AMLs, median 2 mutations per case (range 1 – 6 trackable mutations); Supplementary Figure 1]. Of those (n=319) achieving morphological CR, the smMIPS MRD panel was applicable to 266 (83.4%). MRD assessment could be performed in 201 adult patients of AML (enrolment flow chart in supplementary figure 2). A summary of the clinical and laboratory characteristics of these 201 patients can be seen in Table 1.
  b. *Treatment of AML and MRD Sampling:* All patients were treated with conventional “3+7” induction chemotherapy and further treated with high dose cytarabine (HiDAC) or allogeneic bone marrow transplantation (aBMT), if feasible.^36^ Only 15 patients received aBMT and their outcome was not different from the rest with respect to OS and RFS (p = not significant; supplementary figure 3) and are not considered separately. Sample for FCM-MRD was obtained from the bone marrow at end of induction (PI; n=200) and end of first consolidation cycle (PC, n=98). NGS MRD sample also obtained at the same time points (PI-196; PC-127) from the bone marrow (n=269; PI: 181 and PC:88) or peripheral blood (n=51; PI:15 and PC:36).
  c. *Evaluation of Treatment Outcome:* Overall Survival (OS) and Relapse Free Survival (RFS) were calculated as previously described.^9, 21, 36^ The prognostic impact of NGS and FCM-MRD assays on OS and RFS was computed using the Kaplan-Meier method and compared using log-rank test. Multivariate analysis was performed using the Cox proportional-hazards regression analysis that considered FCM-MRD and NGS-MRD. Separate models were constructed for post induction and post consolidation MRD time points. Grey test was used to compare the cumulative incidences of relapse (CIR) and non-relapse mortality (NRM) using “cmprsk” module in R.^37^ The same module was used to generate representative graphs. Positive predictive value (PPV) and negative predictive value (NPV) were calculated as described.^34^
2. DETECTION OF MRD USING NGS
  a. *smMIPS panel:* We created a 35 gene panel comprising of a pool of 302 smMIPS (as seen in Supplementary Table 1). In brief, this panel covers regions of 35 commonly mutated genes in AML (*ATM, BCOR, DNMT3A, EZH2, FLT3, GATA1, GATA2, IDH1, IDH2, JAK2, KDM6A, KIT, KMT2D, KRAS, NF1, NOTCH1, NOTCH2, NPM1, NRAS, PHF6, PTPN11, RAD21, RUNX1, SETBP1, SF3B1, SH2B3, SMC1A, SRSF2, STAG2, TET2, TP53, U2AF1, WT1, ZRSR2*). The panel was rebalanced (Supplementary Figure 4) to ensure uniform capture across regions. Approximately 600ng of genomic DNA was captured, treated with exonucleases and PCR amplified to create a sequencing ready library. Details pertaining to smMIPS design, assay standardization and sequencing are detailed in supplementary methods.
  b. *Bioinformatics*: Reads were demultiplexed, trimmed, paired end assembled and mapped to the human genome (build hg19). Singleton reads (originating from one UMI) were discarded, and consensus family based variant calling performed using tools described in supplementary methods. We then created a site and mutation specific error model to ascertain the relevance of an observed variant at each site.^35^ Criteria for variant calling using the smMIPS MRD assay are described in supplementary methods.
  c. *MRD for FLT3-internal tandem duplications (ITD): FLT3-ITD* were detected using a novel one-step PCR based NGS assay (see Supplementary Table 3). Variants were detected using a recently described algorithm for the accurate detection of FLT3-ITD.^38^
  d. *Orthogonal detection of MRD in NPM1 mutated AML: NPM1* mutations were additionally tracked using an ultrasensitive orthogonal *NPM1* MRD assay.^21^
3. DETECTION OF MRD IN USING MULTICOLOUR FLOW CYTOMETRY (FCM-MRD) FCM-MRD was detected using a previously described 10 colour two tube MRD assay.^9, 21, 36, 39^ This approach uses a combination of leukemia associated immunophenotype and difference from normal approaches to detect MRD in AML.

**Table 1:**
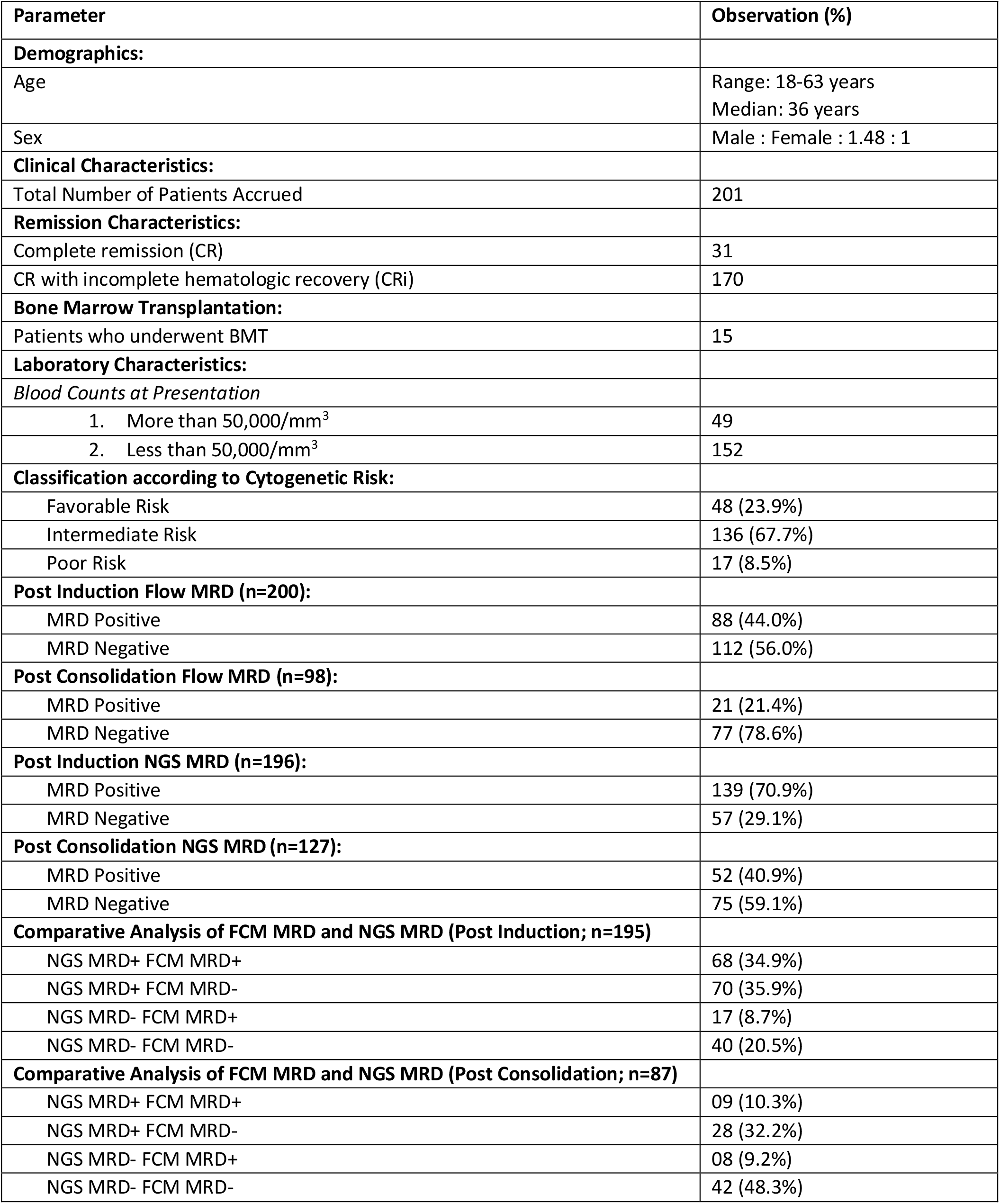
Summary of clinical, laboratory and MRD characteristics of patients accrued in this study.

## Results

1. PATIENTS: The median follow-up of the cohort was 42.3 months. The median OS was 35.9 months (95%CI- 27.2 to 42.8) for the entire cohort. The median RFS was 21.6 months (95%CI- 17.0 to 28.9) months. Additional patient characteristics can be seen in Table 1.
2. NGS BASED AML MRD:
  a. *Assay Characteristics:* We describe an NGS-MRD approach that is applicable to 83.2% of all AML. For patients in morphological CR (n=319) this approach was applicable to 83.4% (n=266). Of these, MRD could be assessed in 201 cases. A co-occurrence plot indicating interactions of mutations tracked by NGS-MRD, prior to therapy, can be seen in Figure 1A. The applicability of this MRD panel, when patients (n=201) are classified by cytogenetic risk is seen in supplementary table 2.
  b. *Limit of Detection:* In a limit of detection experiment (supplementary figure 5), we demonstrated that we could detect leukemic clones till a lower limit of 0.05% (0.03% for *NPM1* mutation). Error modelling of normal samples indicated a higher prevalence of C>T and G>A changes consistent with oxidative DNA damage (supplementary figure 6).^35^ *FLT3-ITD* could be detected at a limit of 0.002% VAF (supplementary figure 7). For smMIPS based MRD, samples were sequenced at a median coverage of 14,728x (11,363x consensus coverage) whereas, for FLT3-MRD assay, the median coverage was 1,396,366x. A total of 344 mutations could be detected in 323 samples (Figure 1B,C) with a median VAF of 0.95% [0.76% after exclusion of mutations in *DNMT3A, TET2, ASXL1* (DTA) genes]. Amongst positive patients, a median of 2 mutations could be detected per patient (range 1-4) at end of induction.
  c. *Clinical Relevance of NGS-MRD:* Nearly 71% (n=139; 70.9%) of patients harboured MRD at the end of induction and 40.9% (n=52) at the end of consolidation. Patients harbouring MRD had a significantly higher CIR as compared to MRD negative patients at PI time point as seen in Figure 2A, Table 2. For patients who were PC NGS-MRD positive a similar observation for CIR was made (Figure 2B, Table 2). The presence of NGS-MRD at the end of induction was associated with inferior OS and RFS (Figure 2C,E) as detailed in Table 3. Similarly, the presence of NGS-MRD (n=52; 40.9%) was predictive of inferior OS and RFS at the end of consolidation (Figure 2D,F; Table 3).
    i. <u>Cytogenetics and NGS-MRD</u>: We did not observe a significant distribution of MRD results when cases were distributed by cytogenetic risk. However, the presence of NGS-MRD was predictive of inferior outcome in intermediate cytogenetic risk (and a tendency in favorable risk) as seen in supplementary figure 8.
    ii. <u>Change in mutations over MRD time points</u>: Amongst paired samples (n=122), 83 samples were positive at PI time point, of which 37 became negative at end of consolidation. For 46 patients in whom MRD persisted a change in MRD profile occurred in 18 patients (39.13%). This included a loss of mutation in most cases (n=14) and gain in the rest (supplementary figure 9). There were 5 patients who were negative at PI time point and became positive at end of consolidation. Of these, relapse was seen in two patients.
    iii. <u>Clinical relevance of the kinetics of NGS-MRD clearance</u>: Patients who cleared NGS-MRD at end of induction (and persisted) had a significantly improved OS [HR- 0.45; 95% CI- 0.22 to 0.9; (p=0.02)] and RFS [HR- 0.49; 95% CI- 0.27 to 0.89; (p=0.01)] as compared to patients who became negative at end of consolidation (supplementary figure 10). Similarly, patients who persistently harboured MRD had a significantly inferior outcome as compared to patients who were MRD negative at both time points (supplementary figure 11). There was no genetic difference observed between these two groups (supplementary figure 12).
  d. *Orthogonal detection of NGS-MRD in NPM1 mutated AML:* Orthogonal comparison of MRD detection in *NPM1* mutated AML was performed in 75 patients (23.2% of all MRD samples; Supplementary Methods, supplementary figure 13). There was a good correlation observed with *NPM1* NGS MRD assay (R^2^=0.71). The PPV of smMIPS MRD assay for *NPM1* mutated AML was 58.5% (95%CI- 52.35 to 64.42%) as compared to a lower PPV of *NPM1* NGS MRD assay [50% (95%CI- 45.34% to 54.66%)]. The *NPM1* NGS MRD assay had a much lower specificity [11.11%(95%CI- 3.11 to 26.06%)] as compared to smMIPS MRD assay for MRD detection in *NPM1* mutated AML [46.58%(95%CI- 34.80 to 58.63%)].
3. FCM BASED AML-MRD: The presence of FCM-MRD was associated with inferior OS, RFS and CIR at the end of induction and consolidation as detailed in Tables 2, 3 and supplementary figure 14.
4. COMPARISON OF FCM AND NGS MRD: On incorporating results combining both the MRD modalities, patients that were positive by both techniques (FCM+ NGS+) had a significantly inferior outcome with respect to OS, RFS and CIR at any MRD time point as compared to patients negative by both modalities as seen in Table 2,3 (Figure 3). A comparison of the baseline mutational profiles between dual PI MRD positive and negative groups revealed a significantly higher (p=0.04) prevalence of *RUNX1* mutations in the dual MRD positive subset (supplementary figure 15).
  a. *Clinico-pathological correlates of FCM MRD Positive NGS MRD Negative AML*: A total of 20 patients were (FCM+NGS-) at assessed at PI and/or PC time points, of which 4 were detected at subsequent (or preceding) MRD time point. Of the remaining 16, in three patients a paired MRD sample was lacking. Amongst the rest, six patients (out of 13) had relapsed, three died due to non-relapse causes and four were alive at last follow up (supplementary table 5).
  b. *Accuracy of NGS-MRD in comparison with FCM-MRD:* The PPV and NPV metrics of end of induction NGS-MRD to predict relapse in AML were 70.5% and 57.89% respectively with an accuracy of 66.84%. FCM MRD metrics at end of induction were comparable for PPV (75%), NPV (48.2%) and accuracy to predict relapse(60%). At the PI time point, NGS-MRD correctly identified 80% (68 out of 85) of cases classified as MRD positive by FCM, whereas FCM identified just 68 out of 138 cases (49.3%) identified by NGS. A detailed comparison of PPV, NPM and accuracy of combinations of patients detected between these two assays can be seen in supplementary table 6.
5. MULTIVARIATE ANALYSIS: Both FCM and NGS MRD were important in predicting outcome as seen in Table 3. However, PI NGS-MRD emerged as the most important independent prognostic factor predictive of inferior outcome for OS [HR- 1.94; 95% CI- 1.15 to 3.27; (p<0.0001)] as well as RFS [HR- 2.05; 95% CI- 1.30 to 3.23; (p<0.0001)].

**Figure 1:**
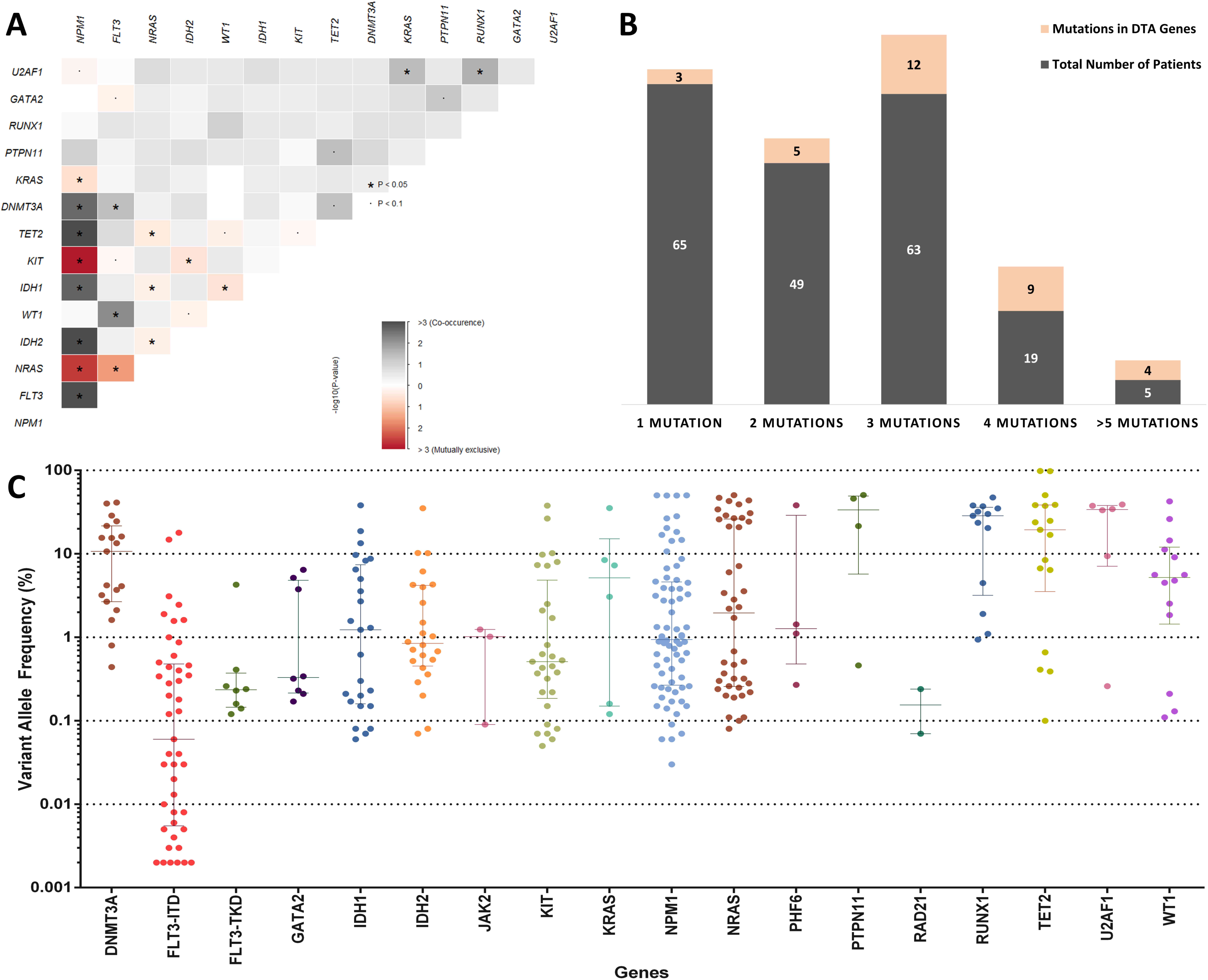
Somatic mutations in AML detected at diagnosis and during therapy. Figure 1A: The interaction of mutations at baseline is demonstrated here using Fisher’s Exact test. Co-occurrence is indicated in grey colour and mutual exclusivity is indicated in red. Figure 1B: The total number of mutations detected per patient and the number of such patients in the cohort is displayed. The total number of mutations in *DNMT3A-TET2-ASXL1* genes is indicated here as a fraction. Figure 1C: Variant allele frequencies of mutations detected at MRD time points for patients of AML in morphological remission. The bars indicate median values with interquartile ranges.

**Figure 2:**
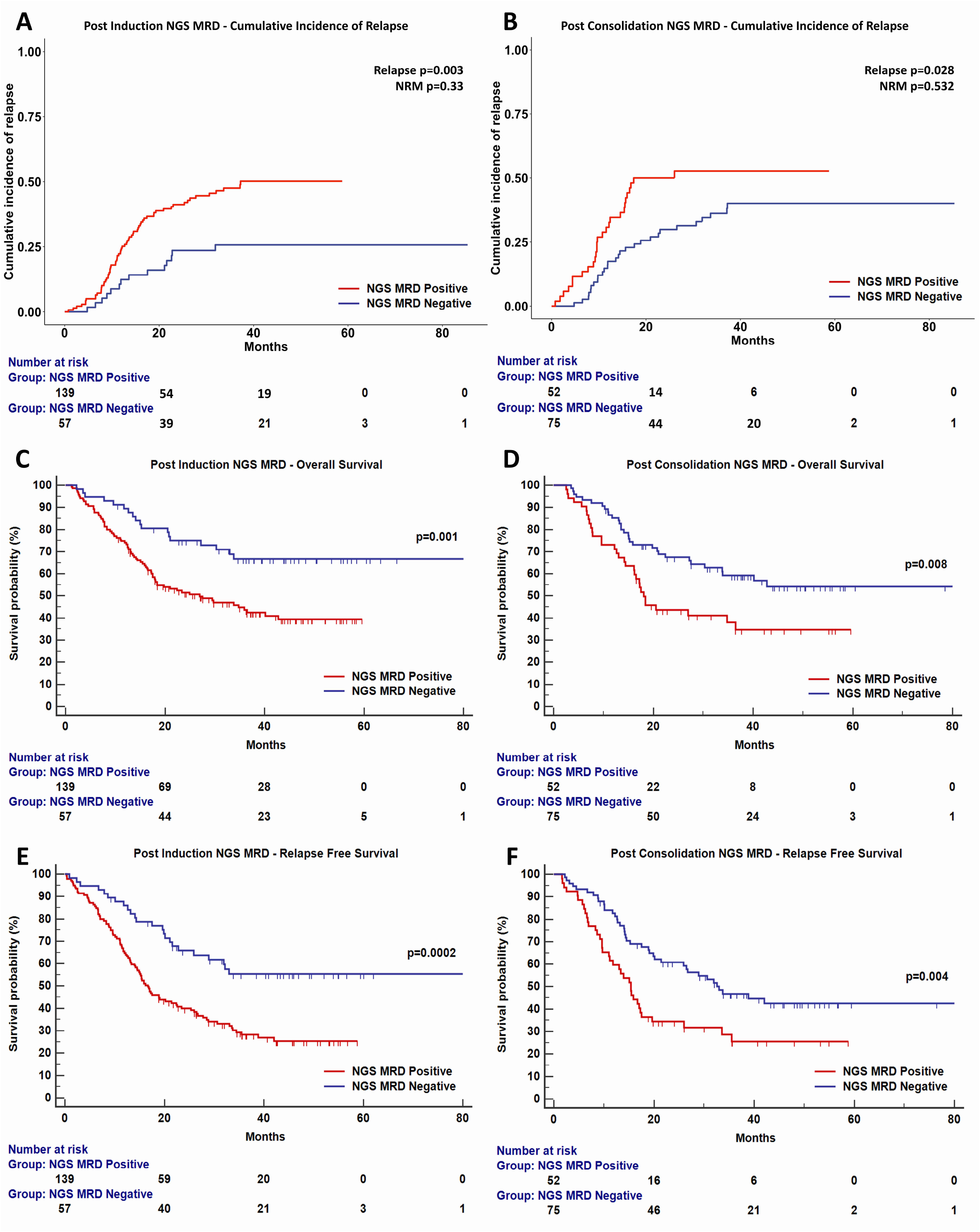
Clinical relevance of error corrected NGS-MRD. Presence of NGS MRD at post induction (A) and post consolidation time points (B) is associated with a higher cumulative incidence of relapse (CIR). Kaplan Meyer plots indicate the clinical relevance of NGS-MRD with respect to OS and RFS at post induction (C,E) and post consolidation time points (D,F).

**Figure 3:**
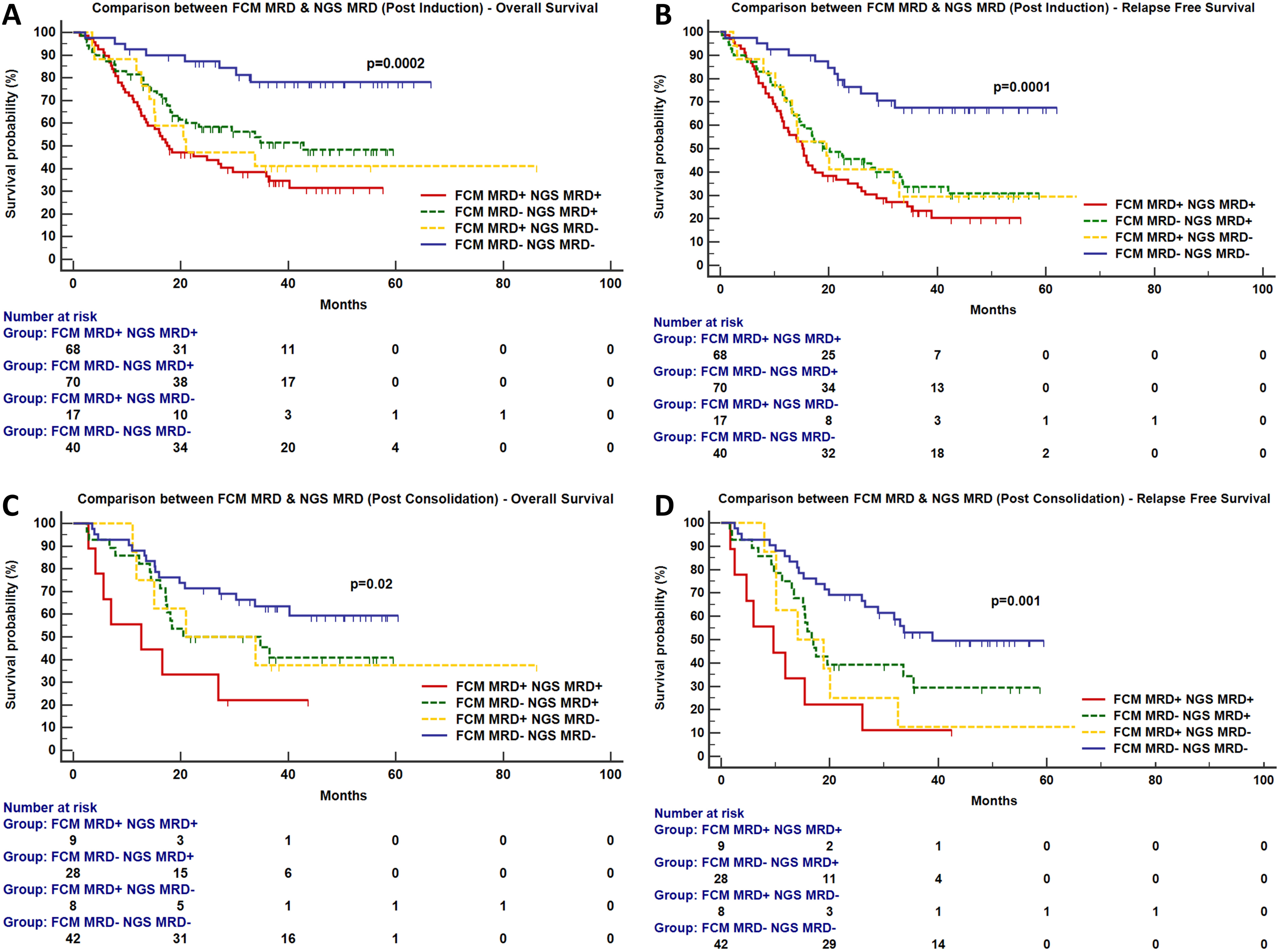
Comparison between FCM and NGS-MRD. The clinical relevance detection of MRD during complete remission when measured by FCM or error corrected NGS at post induction (A,B) and post consolidation time points (C,D).

**Table 2:** Prognostic influence of NGS-MRD, FCM-MRD and a combination of these modalities on the cumulative incidence of relapse (CIR)

| Cumulative Incidence of Relapse |  |  |  |  |  |  |
| --- | --- | --- | --- | --- | --- | --- |
| Post Induction NGS MRD | Cumulative Incidence of Relapse (CIR) |  |  | Non-Relapse Mortality (NRM) |  |  |
|  | HR | 3-year CIR (95% CI) | P-value | HR | 3-year Cumulative Incidence (95% CI) | P-value |
| MRD Negative | 1 | 25.7 % (14.9 to 37.9) | 0.003 | 1 | 18.7 % (9.5 to 30.3) | 0.333 |
| MRD Positive | 8.81 | 47.5 % (38.7 to 55.8) |  | 0.93 | 23.4 % (16.6 to 30.8) |  |
| Post Consolidation NGS MRD | Cumulative Incidence of Relapse |  |  | Non-Relapse Mortality |  |  |
|  | HR | 3-year CIR (95% CI) | P-value | HR | 3-year Cumulative Incidence (95% CI) | P-value |
| MRD Negative | 1 | 36.2 % (25.1 to 47.4) | 0.03 | 1 | 16.6 % (9.0 to 26.1) | 0.532 |
| MRD Positive | 4.79 | 52.8 % (37.8 to 65.7) |  | 0.39 | 19.2 % (9.8 to 31.0) |  |
| Post Induction FCM MRD | Cumulative Incidence of Relapse |  |  | Non-Relapse Mortality |  |  |
|  | HR | 3-year CIR (95% CI) | P-value | HR | 3-year Cumulative Incidence (95% CI) | P-value |
| MRD Negative | 1 | 33.6 % (24.7 to 42.7) | 0.008 | 1 | 19.8 % (12.7 to 28.0) | 0.321 |
| MRD Positive | 6.97 | 49.8 % (38.6 to 59.9) |  | 0.98 | 25.2 % (16.6 to 34.8) |  |
| Post Consolidation FCM MRD | Cumulative Incidence of Relapse |  |  | Non-Relapse Mortality |  |  |
|  | HR | 3-year CIR (95% CI) | P-value | HR | 3-year Cumulative Incidence (95% CI) | P-value |
| MRD Negative | 1 | 35.9 % (25.1 to 46.9) | 0.01 | 1 | 19.9 % (11.7 to 29.7) | 0.632 |
| MRD Positive | 6.03 | 61.9 % (36.3 to 79.7) |  | 0.22 | 23.8 % (8.2 to 43.9) |  |
| Comparative Analysis of FCM MRD and NGS MRD (Post Induction) | Cumulative Incidence of Relapse |  |  | Non-Relapse Mortality |  |  |
|  | HR | 3-year CIR (95% CI) | P-value | HR | 3-year Cumulative Incidence (95% CI) | P-value |
| NGS MRD- FCM MRD- | 1 | 18.7 % (8.0 to 32.7) | 0.003 | 1 | 13.8 % (4.8 to 27.3) | 0.425 |
| NGS MRD+ FCM MRD- | - | 41.9 % (29.6 to 53.7) |  | - | 23.7 % (14.2 to 34.6) |  |
| NGS MRD- FCM MRD+ | - | 41.2 % (17.5 to 63.6) |  | - | 29.4 % (9.8 to 52.4) |  |
| NGS MRD+ FCM MRD+ | 14.01 | 52.5 % (39.6 to 63.9) |  | 2.78 | 23.5 % (14.2 to 34.2) |  |
| Comparative Analysis of FCM MRD and NGS MRD (Post Consolidation) | Cumulative Incidence of Relapse |  |  | Non-Relapse Mortality |  |  |
|  | HR | 3-year CIR (95% CI) | P-value | HR | 3-year Cumulative Incidence (95% CI) | P-value |
| NGS MRD- FCM MRD- | 1 | 24.8 % (12.6 to 39.0) | 0.0003 | 1 | 22.0 % (10.7 to 35.9) | 0.398 |
| NGS MRD+ FCM MRD- | - | 50.0 % (29.9 to 67.1) |  | - | 17.8 % (6.3 to 34.1) |  |
| NGS MRD- FCM MRD+ | - | 87.5 % (17.2 to 98.9) |  | - | 0.0 % (-) |  |
| NGS MRD+ FCM MRD+ | 18.81 | 55.5 % (14.7 to 83.5) |  | 2.95 | 33.3 % (5.7 to 65.6) |  |
| Dual Timepoint NGS MRD | Cumulative Incidence of Relapse |  |  | Non-Relapse Mortality |  |  |
|  | HR | 3-year CIR (95% CI) | P-value | HR | 3-year Cumulative Incidence (95% CI) | P-value |
| MRD Negative | 1 | 27.8 % (13.5 to 44.0) | 0.04 | 1 | 15.9 % (5.6 to 31.1) | 0.834 |
| Either MRD Positive | - | 44.7 % (28.6 to 59.7) |  | - | 19.0 % (8.8 to 32.2) |  |
| MRD Positive | 6.46 | 52.9 % (37.0 to 66.6) |  | 0.36 | 17.4 % (8.0 to 29.7) |  |

**Table 3:**
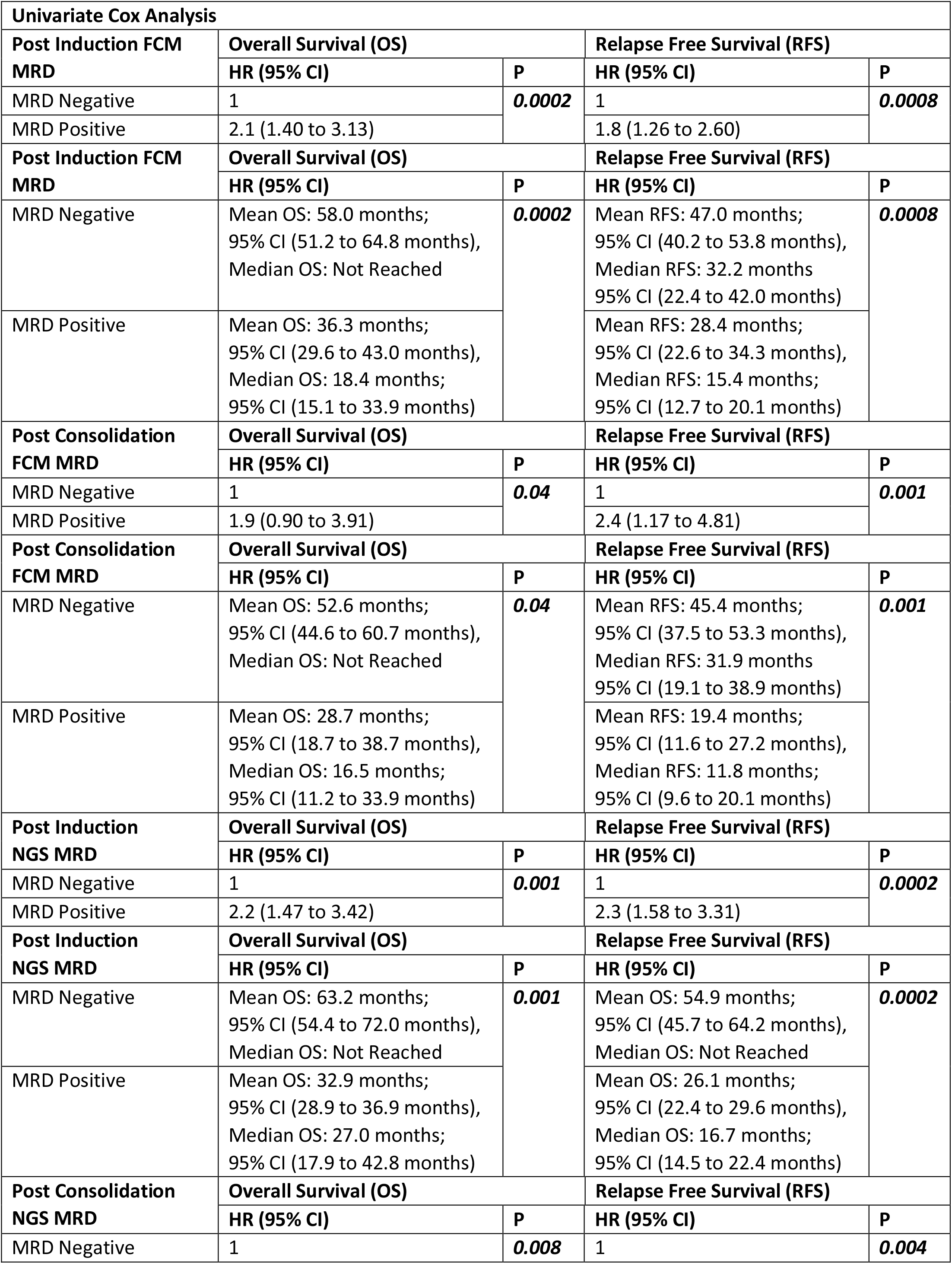
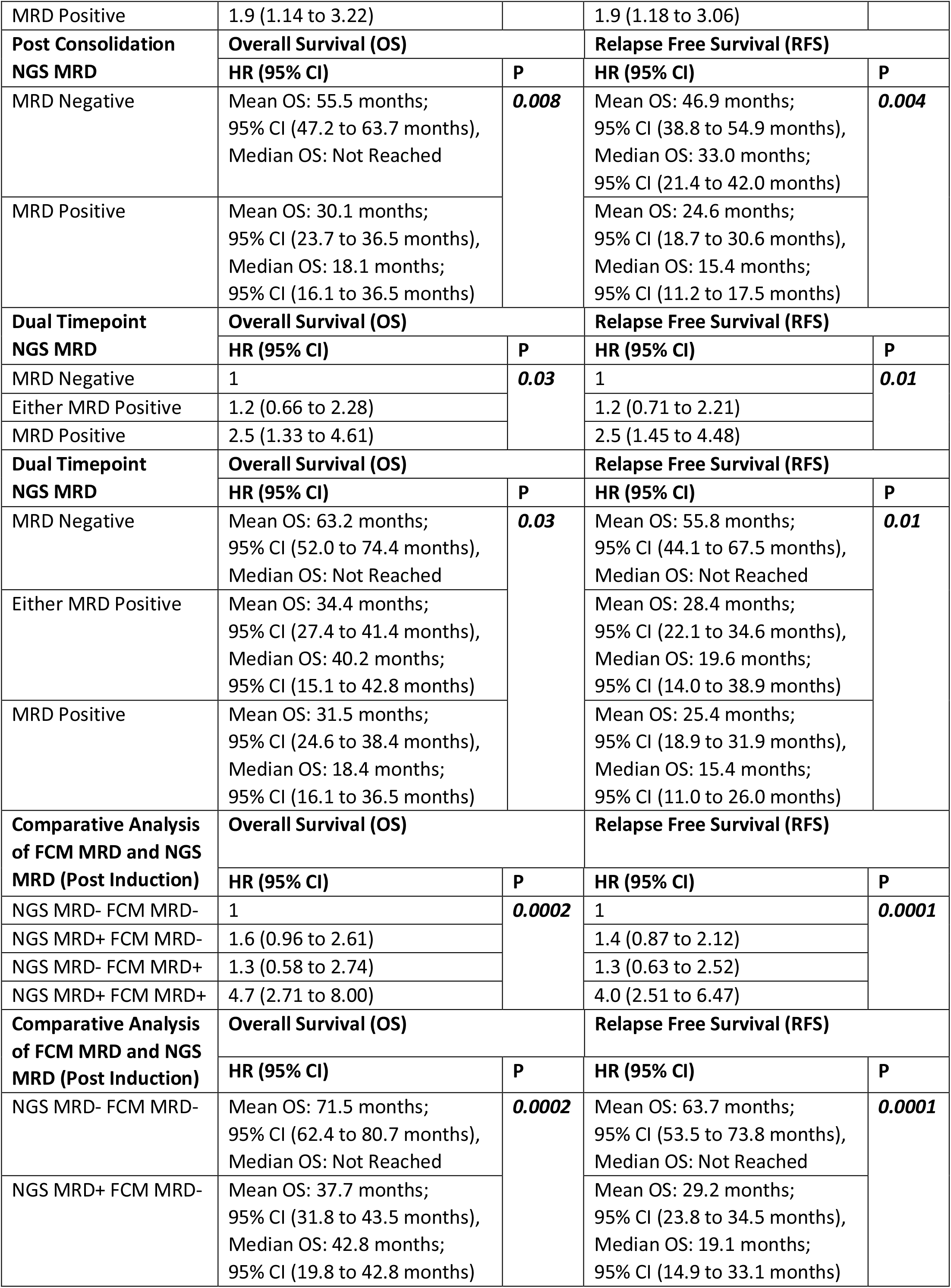
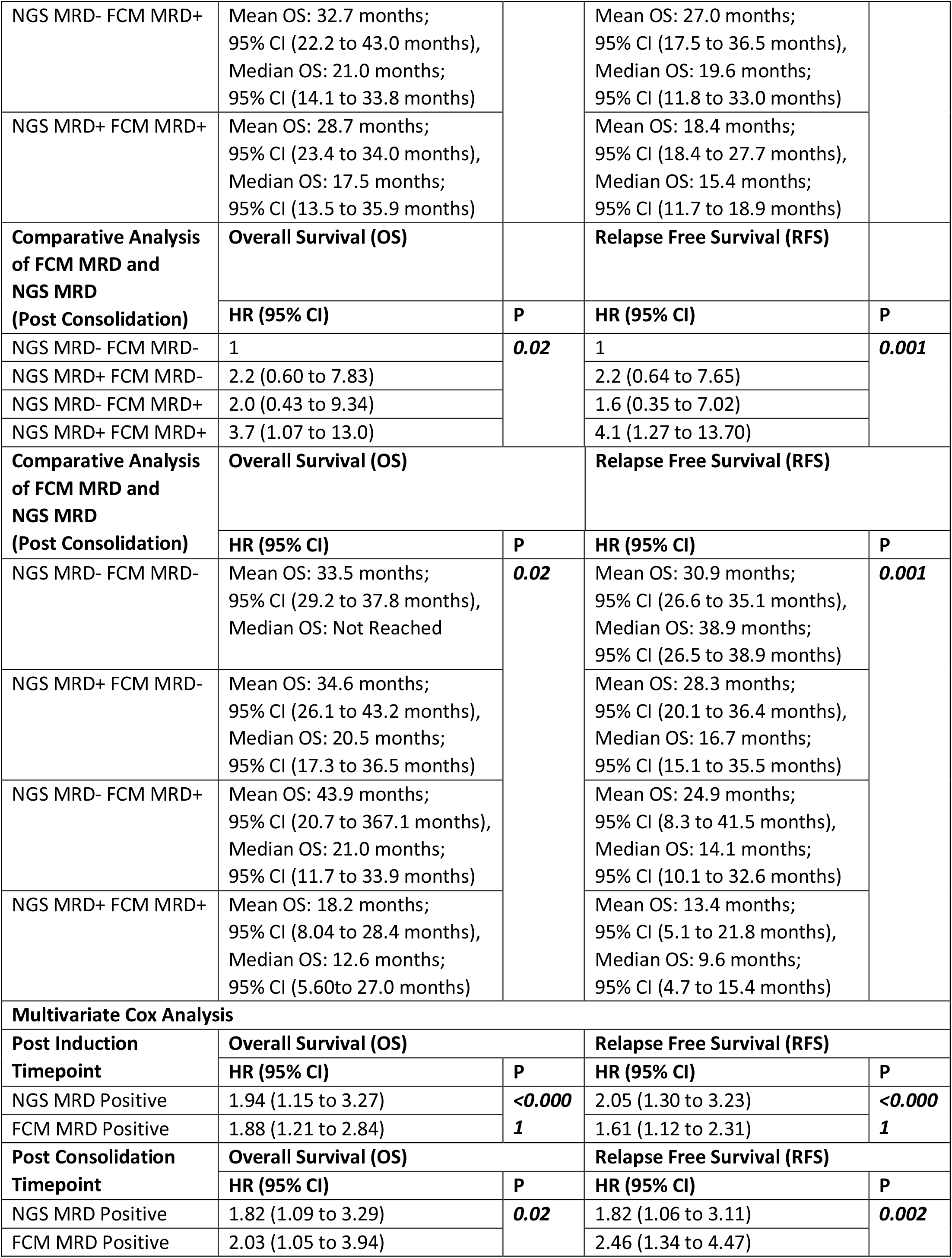
Difference in OS and RFS between FCM-MRD, NGS-MRD and comparative analysis between the two modalities at post Induction and post consolidation timepoints. OS: Overall Survival, RFS: Relapse Free Survival, CI: confidence interval.

## Discussion

Recently, Hourigan and colleagues ^40^ performed ultradeep sequencing using a 13-gene panel to detect MRD in AML. In a pioneering effort, they demonstrated an advantage of myeloablative conditioning in preventing relapse in an AML cohort based on NGS MRD results. The authors, however, were unable to compare their results with other MRD assessment techniques. Here, we have assessed MRD at serial time points and have compared our results with 10 colour FCM-MRD, which is a widely used technique for the assessment of response to chemotherapy.^41^ We find that NGS-MRD is comparable in applicability and adds value, especially when a clear distinction of regenerating myeloid progenitors from leukemic blasts is absent. Advantages over FCM-MRD include potential for multi-institutional standardization, an ability to scale up operations and lack of requirement of expert operators. In our manuscript we demonstrate that NGS-MRD identified over 80% of cases identified by flow cytometry at PI time point. On evaluating 20 cases (from both PI and PC timepoints) missed by NGS-MRD (but detected by FCM), we observed that 20% of these were detected by NGS-MRD at a subsequent or preceding MRD time point. An additional 56.3% (n=9) are alive or have died due to causes other than relapse indicating that these could have been false positives. In that case, only a fraction of cases (n=7) are truly missed by NGS as compared to FCM-MRD. An additional novel insight gained is gained by studying the kinetics of MRD (supplementary figure 10). We demonstrate that patients who became negative at end of induction are likely to have a superior outcome as patients who turn negative at a subsequent time point.

Previously Jongen-Lavrencic have demonstrated clinical utility of NGS to detect MRD in AML by using computational error correction to mitigate sequencing errors.^27^ Such an approach although easy to implement discounts for batch effects and variability that occurs because of library clustering and batch dependent PCR artefacts.^32^ In that context, to the best of our knowledge, this is the first study to determine the clinical importance of (error corrected, panel based) NGS-MRD in AML treated with conventional therapies. Although our NGS-MRD strategy works in a majority of AML, we were curious about the genetic basis of cases (n=65 out of 393) in which this strategy *did not* work. The cytogenetic and mutational landscape can be seen in supplementary figure 16. Nearly half of these patients did not harbour any mutation at diagnosis (n=29; 44.6%). Insight into rest of the cases revealed *ASXL2* as a recurrently mutated gene (n=8,12.3%). Incorporation of *ASXL2* in future iterations of our panel will as well as other UMI based RNA sequencing approaches^24,25^ to monitor chimeric gene fusions will help in increasing the breadth of our approach.

Consistent with previous reports, we find that in some patients, mutations in DTA genes are present at high VAF at MRD time points (Figure 1) indicating an origin from an ancestral clone possibly originating from clonal haematopoesis.^27, 34, 40^ We are in agreement with Thol and colleagues who indicated that mutations in *TET2* could reliably be used as MRD markers, at least in a fraction of cases. ^34^ Unlike amplicon based^28,34^ approaches, the advantage of a smMIPS based capture includes a stable panel which can be used across a spectrum of cases and no susceptibility to allelic skew or PCR induced errors prior to incorporation of the UMI barcode. Disadvantages of smMIPs include poor performance for GC rich genes such as *CEBPA* gene and inability to capture low yield or poor quality of DNA (a problem not infrequently seen with MRD samples). The library preparation process, is relatively low cost in nature and the overall process has a realistic, turnaround time of five to seven days. Our observation is that sensitivity in the clinic for most mutations is close to 0.1% VAF. A lower can be obtained for complex indels such as *NPM1* and *FLT3-ITD*. Improvements with sensitivity may be possible through duplex sequencing based methods albeit at a much higher cost of sequencing. ^30^ Lastly, based on this data we find that mutations in *NPM1, FLT3, NRAS, KIT, IDH1, IDH2, WT1, RUNX1, GATA2, U2AF1* and *PHF6* were most helpful in making an positive NGS-MRD call. (supplementary figure 17).

To conclude, we demonstrate that panel based error corrected NGS-MRD is clinically relevant and possibly advantageous when compared to FCM-MRD based AML MRD.

## Data Availability

Agreement to Share Publication-Related Data and Data Sharing Statement:
Dr Nikhil Patkar, Haematopathology Laboratory, CCE Building, Advanced Centre for Treatment Research and Education in Cancer, Tata Memorial Centre, Kharghar, Maharashtra, India, Pin: 410210. Phone: +91-22-27405000, Ext: 5773

## Acknowledgements

This work was supported by the Wellcome Trust/DBT India Alliance Fellowship [grant number IA/CPHI/14/1/501485] awarded to Dr Nikhil Patkar.

We are grateful for the training imparted to Dr Nikhil Patkar in the field genome sequencing technologies by Dr David Wu, Dr Stephen Salipante and Dr Brent Wood in the Department of Laboratory Medicine, University of Washington, USA & Salipante Lab, University of Washington. We are greatful to Dr Jay Shendure (Department of Genome Sciences, University of Washington) for agreeing to be an the external sponsor for Wellcome Trust/DBT India Alliance Fellowship. We are also greatful to Dr Christopher Hourigan for critically reviewing this manuscript and providing very helpful comments and suggestions.

## Authorship and conflict-of-interest statements

NP designed the study, conducted research, analyzed, interpreted the data, and wrote the manuscript. AFS, CK, RS, PB, SR, SJ, SC, RK conducted research conducted research and analyzed data. DS, SG, GC, ND, PT, PGS, SG conducted research and analyzed data. BB, HM, NK, MS recruited patients and analyzed data.

The author(s) declare no competing financial interests.

## Agreement to Share Publication-Related Data and Data Sharing Statement

Dr Nikhil Patkar, Haematopathology Laboratory, CCE Building, Advanced Centre for Treatment Research and Education in Cancer, Tata Memorial Centre, Kharghar, Maharashtra, India, Pin: 410210. Phone: +91-22-27405000, Ext: 5773

## Supplementary Methods

### Patkar et al. Molecular MRD Detection in AML using Error Corrected NGS

#### 1. Detection of AML MRD using smMIPS

**Supplementary Figure 1:**
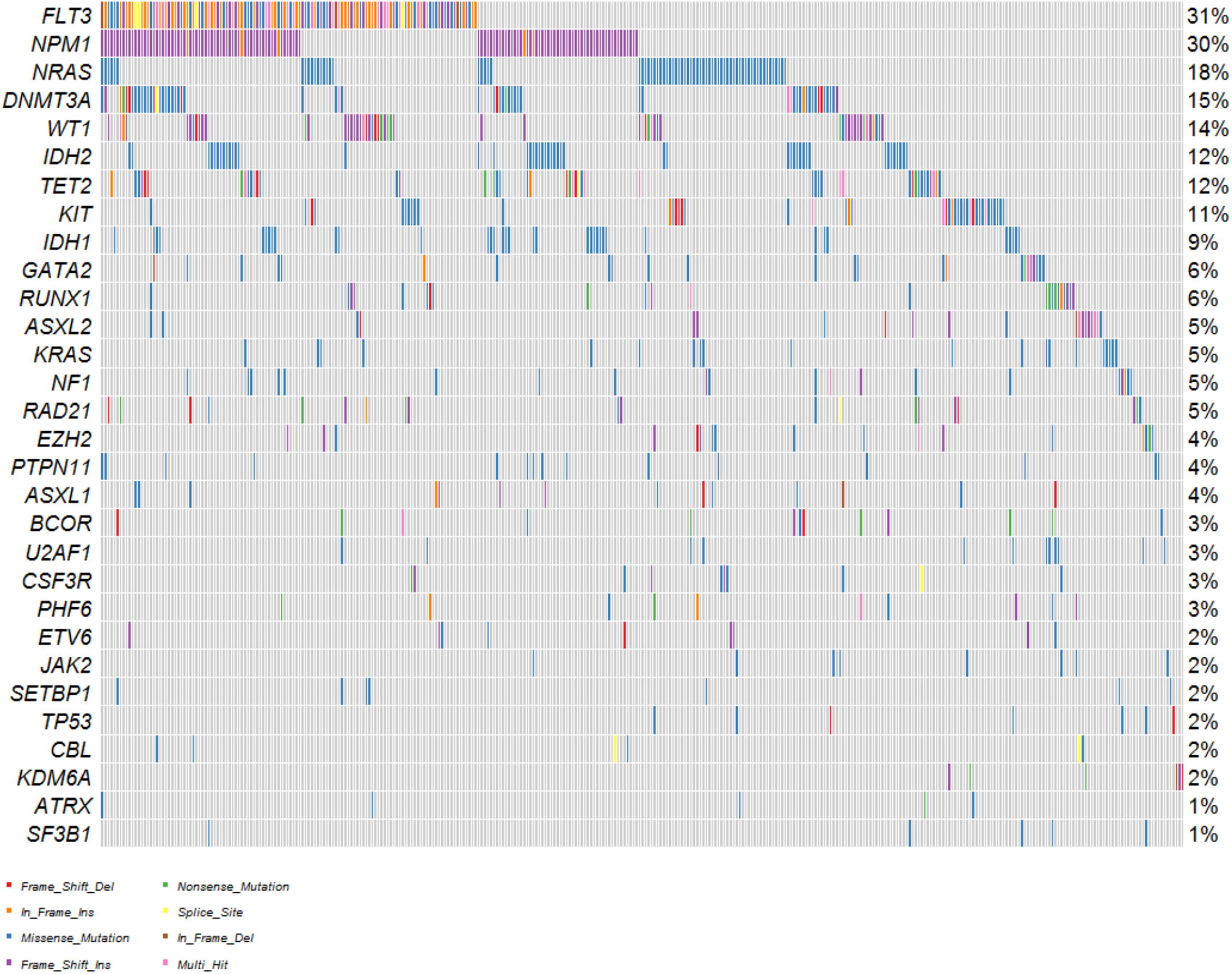
Above oncoplot (on 393 patients) demonstrates the frequency of somatic mutations at diagnosis in AML detected using a 50 gene smMIPS myeloid panel. This genomic landscape formed the basis for designing the NGS MRD panel.

##### a. Design of smMIPS

We reviewed somatic mutations in 393 patients of adult AML (Supplementary Figure 1) diagnosed at the Tata Memorial Centre as well as mutations in TCGA AML cohort (www.cbioportal.org). Based on this data we designed a 35 gene myeloid hotspot panel comprising of 302 single molecule molecular inversion probes (smMIPS). The genes covered by this “hot-spot” panel can be seen in Supplementary Table 1. The panel comprised of a main panel and an add on module which covered uncommon mutations. The latter panel was added if mutations (covered by addon panel) were present at diagnosis. smMIPS were designed using MIPgen^1^ software with the parameters -max_capture_size 162, -min_capture_size 152, -logistic_priority_score 0.5. Extenstion arm length parameters were set between 18-21 and ligation arm length between 21-24. Each smMIP was designed to include a four basepair (bp) unique molecular identifier (UMI) at each end (total of 8bp degenerate nucleotide sequence per smMIP).

**Supplementary Figure 2:**
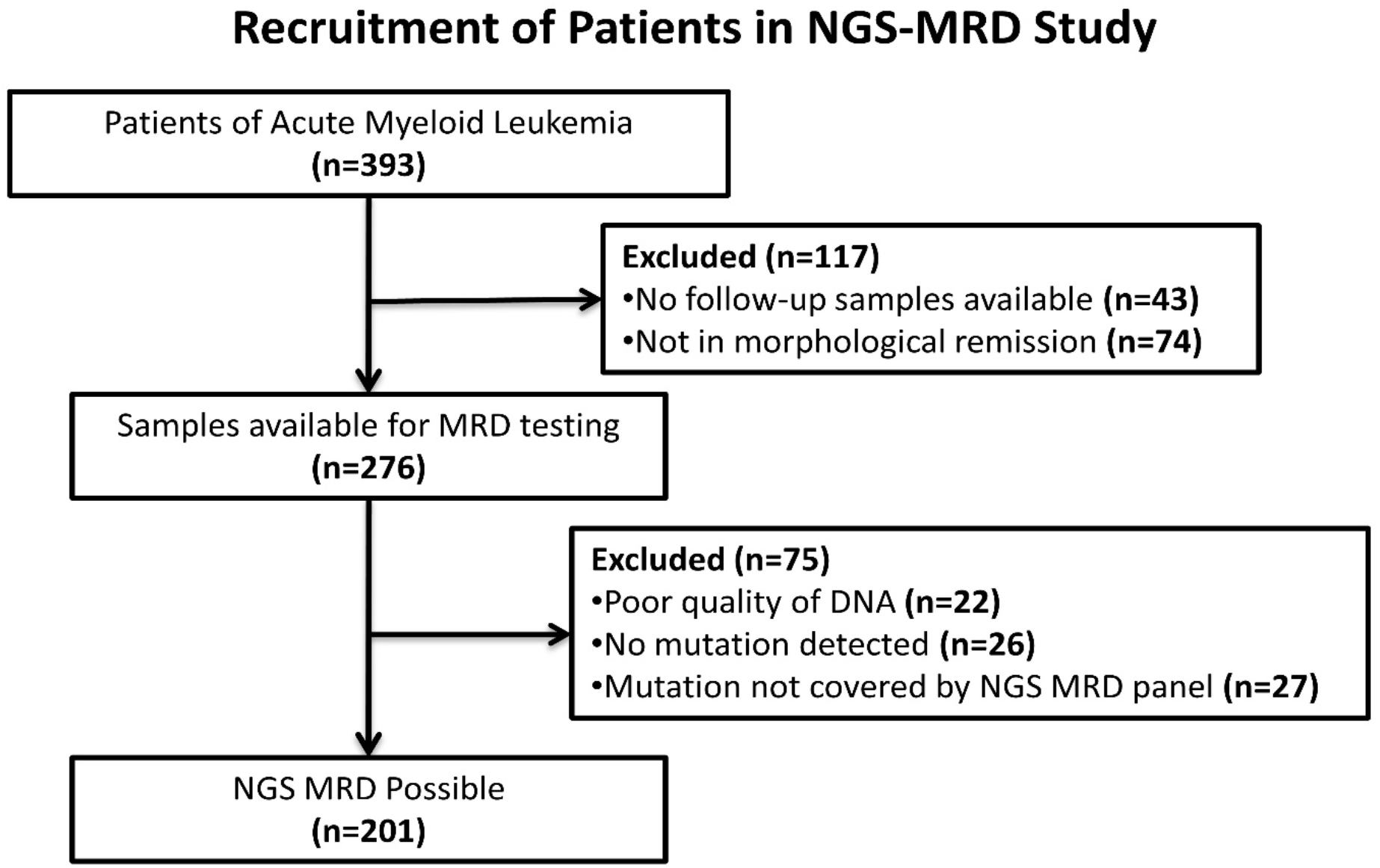
Flow chart indicates recruitment of patients in AML NGS-MRD study.

**Supplementary Figure 3:**
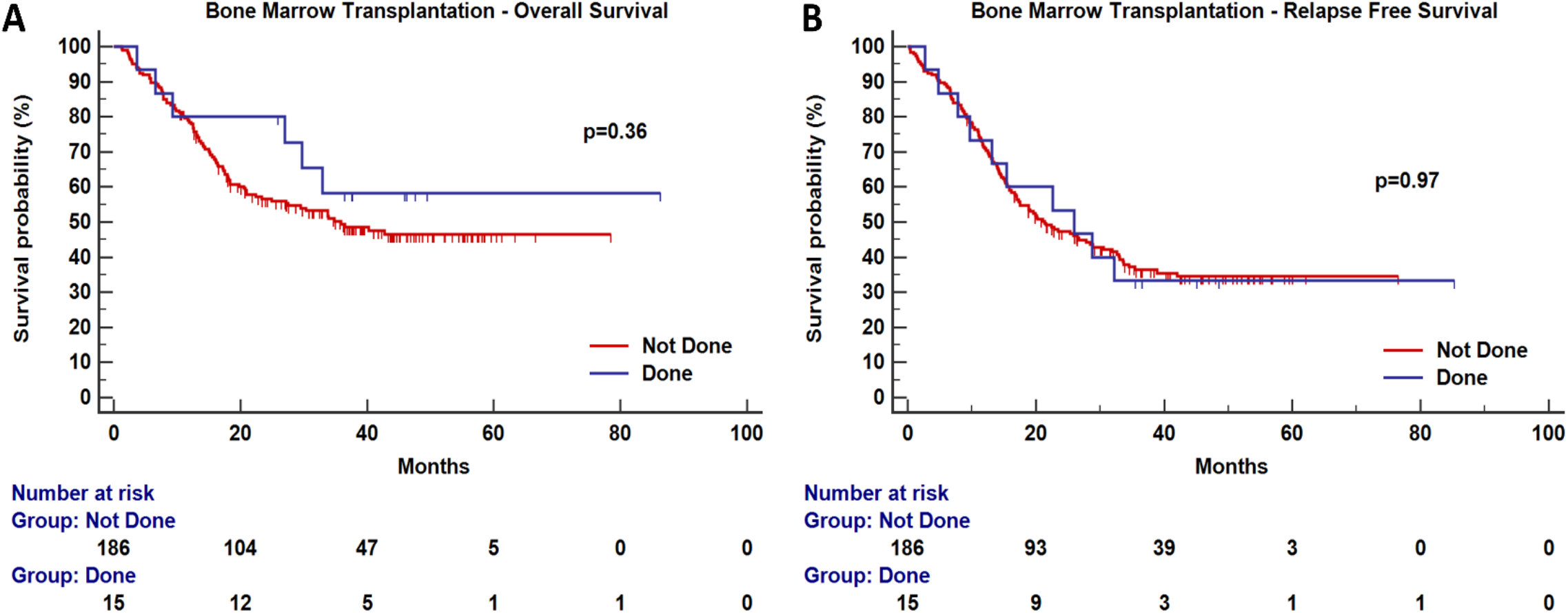
Survival estimates for overall survival (A) and relapse free survival (B) of allogeneic bone marrow transplantation in this patient cohort.

##### b. Rebalancing smMIPS to ensure uniform capture

Following initial equimolar pooling and analysis of read depths, the smMIPS pool underwent two rounds of rebalancing. A balanced version can be seen in Supplementary Figure 2.

**Supplementary Figure 4:**
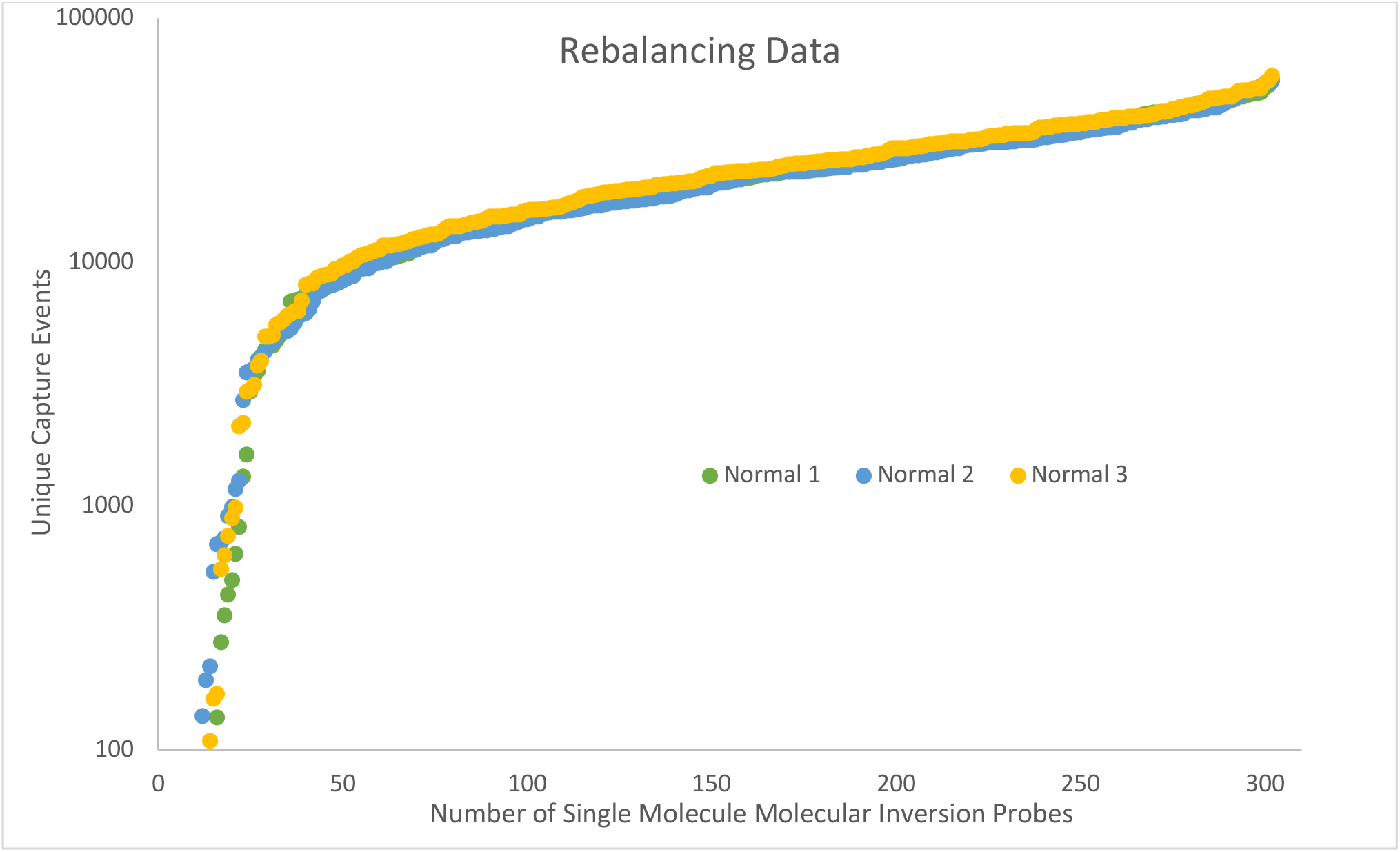
Unique Capture Events per smMIP for three normal specimens.

##### a. Sequencing

Initial standardization and balancing experiments using NA12878 control DNA were performed on a MiSeq (standard V2 flow cell 150PE chemistry). Once the assay was standardized, error modelling, limit of detection and MRD detection experiments were performed on multiple S4 flow cells of a NovaSeq 6000 using 150PE chemistry.

##### b. Data Analysis

The bioinformatics approach was similar to that published by Waalkes et al^2^ with a few modifications. Demultiplexing was performed using bcl2fastq-v2.17. Adapter sequences, smMIPs backbone and reads less than 53bp were trimmed using fastq-mcf tool of ea-utils (https://expressionanalvsis.github.io/ea-utils/). Paired end assembly was carried out using PEAR (vO.9.8).^3^ A custom script was used to trim, concatenate the 4bp UMI at the 5’ and 3’ end of assembled read and add it to the read header. Reads were mapped to the human genome (build hg19) using bwa-0.7.12^4^ and pre-processed using SAMtools-1.1^5^. For computational efficiency, mapped reads were split by chromosome and into files sourced from reads mapping to individual smMIPS based on their genomic start and stop coordinates. Each of these files is processed individually for downstream variant calling. All reads originating from a single UMI were discarded (singleton reads). The rest of the reads (two or more UMI) were used to create a consensus .sam file using CallMolecularConsensusReads function of fgbio-0.4.0 with the following parameters --error-rate-post-umi=30 --min-reads=2 --min-input-base-quality 20 (https://github.com/fulcrumgenomics/fgbio). These reads were converted to fastq using picard-2.17.2 (http://broadinstitute.github.io/picard/) and mapped, sorted and indexed using bwa and samtools as described above. A .mpileup was created using samtools-1.1 and sequence variants detected using a bespoke variant caller. (https://bitbucket.org/uwlabmed/smmips_analysis/src/master/). Variant calls from individual smMIPs were combined into a single .vcf file and annotated using ANNOVAR.^6^

**Supplementary Table 1:** List of genes and their loci sequenced using the smMIPS AML MRD panel. *FLT3-ITD MRD was performed using a one-step PCR based ultradeep sequencing assay

| AML NGS MRD Panel |  |  |  |
| --- | --- | --- | --- |
| Sr. Nos | Gene Name | Region | RefSeqID |
| 1 | *ASXL1* | Exon 13 | NM_015338 |
| 2 | *ATM* | Exon 37 | NM_000051 |
| 3 | *BCOR* | Exon 4, 10, 11 | NM_001123383 |
| 4 | *DNMT3A* | Exon 23 | NM_022552 |
| 5 | *EZH2* | Exon 14-18 | NM_152998.2 |
| 6 | *FLT3* | Exon 20, 14-15* | NM_004119 |
| 7 | *GATA1* | Exon 2 | NM_002049 |
| 8 | *GATA2* | Exon 3, 4 | NM_032638 |
| 9 | *IDH1* | Exon 4 | NM_005896 |
| 10 | *IDH2* | Exon 4, Exon 4 (R172K) | NM_002168 |
| 11 | *JAK2* | Exon 14 | NM_004972 |
| 12 | *KDM6A* | Exon 17, 20 | NM_021140 |
| 13 | *KIT* | Exon 8-11, 17 | NM_000222 |
| 14 | *KMT2D* | Exon 34, 39 | NM_003482 |
| 15 | *KRAS* | Exon 2-4, 5 | NM_004985 |
| 16 | *NF1* | Exon 24, 47 | NM_001042492 |
| 17 | *NOTCH1* | Exon 27, 34 | NM_017617 |
| 18 | *NOTCH2* | Exon 34 | NM_024408 |
| 19 | *NPM1* | Exon 11 | NM_002520 |
| 20 | *NRAS* | Exon 2-3 | NM_002524 |
| 21 | *PHF6* | Exon 2, 3-9 | NM_032458 |
| 22 | *PTPN11* | Exon 3, 13-14 | NM_002834 |
| 23 | *RAD21* | Exon 5, 11 | NM_006265 |
| 24 | *RUNX1* | Exon 4-8 | NM_001754 |
| 25 | *SETBP1* | Exon 4 | NM_015559 |
| 26 | *SF3B1* | Exon 14-16 | NM_012433 |
| 27 | *SH2B3* | Exon 3, 6 | NM_005475 |
| 28 | *SMC1A* | Exon 2, 11 | NM_006306 |
| 29 | *SRSF2* | Exon 1 | NM_003016 |
| 30 | *STAG2* | Exon 7-8, 19, 24, 27-29 | NM_006603 |
| 31 | *TET2* | Exon 3-11 | NM_001127208 |
| 32 | *TP53* | All Exons | NM_001126114 |
| 33 | *U2AF1* | Exon 2, 6, 7 | NM_006758 |
| 34 | *WT1* | Exon 6-8, 11 | NM_024426 |
| 35 | *ZRSR2* | Exon 10 | NM_005089 |

**Supplementary Table 2:** Applicability of NGS-MRD panel in AML patients stratified by cytogenetic risk and number of mutations seen in DTA genes

|  | Favourable Risk |  | Intermediate Risk |  | Poor Risk |  |
| --- | --- | --- | --- | --- | --- | --- |
|  | Total Number of Patients | Mutations in DTA Genes | Total Number of Patients | Mutations in DTA Genes | Total Number of Patients | Mutations in DTA Genes |
| **1 Mutation** | 26 | 2 | 32 | 1 | 7 | 0 |
| **2 Mutations** | 13 | 0 | 33 | 5 | 3 | 0 |
| **3 Mutations** | 9 | 1 | 48 | 11 | 6 | 0 |
| **4 Mutations** | 0 | 0 | 18 | 9 | 1 | 0 |
| **>5 Mutations** | 0 | 0 | 5 | 4 | 0 | 0 |

##### c. Limit of Detection Experiment

In order to determine the limit of detection, we performed an experiment where OCIAML3 cell line [harbouring *DNMT3A* (p.R882C), *NRAS* (p.Q61L) and *NPM1* (Type A mutation)] was serially diluted in a normal bone marrow (BM). Similarly, baseline DNA from five AML samples [each harbouring *NRAS* (p.G12D), *IDH1* (p.R132G), *IDH2* (p.R140Q), *IDH2* (p.R172K) and *NPM1* (type A)] were serially diluted in normal BM as seen in Supplementary Figure 3. The expected variant allele frequency (VAF) for that mutation was calculated from the original VAF found in undiluted sample. The range of the expected VAFs was from 1.25% to 0.02%. We could successfully detect VAFs in nearly all cases up to a lower limit of approximately 0.05%. The limit of detection of NGS-MRD assay was thus at 0.05% for all mutations and 0.03% for the *NPM1* mutation. A lower LOD threshold was acceptable for *NPM1* mutation because of the uniqueness of the indel mutation and a previous observation (based on a limit of blank study) that complex 4bp indels (seen in *NPM1)* are not observed for short read sequencing data.^7^

**Supplementary Figure 5:**
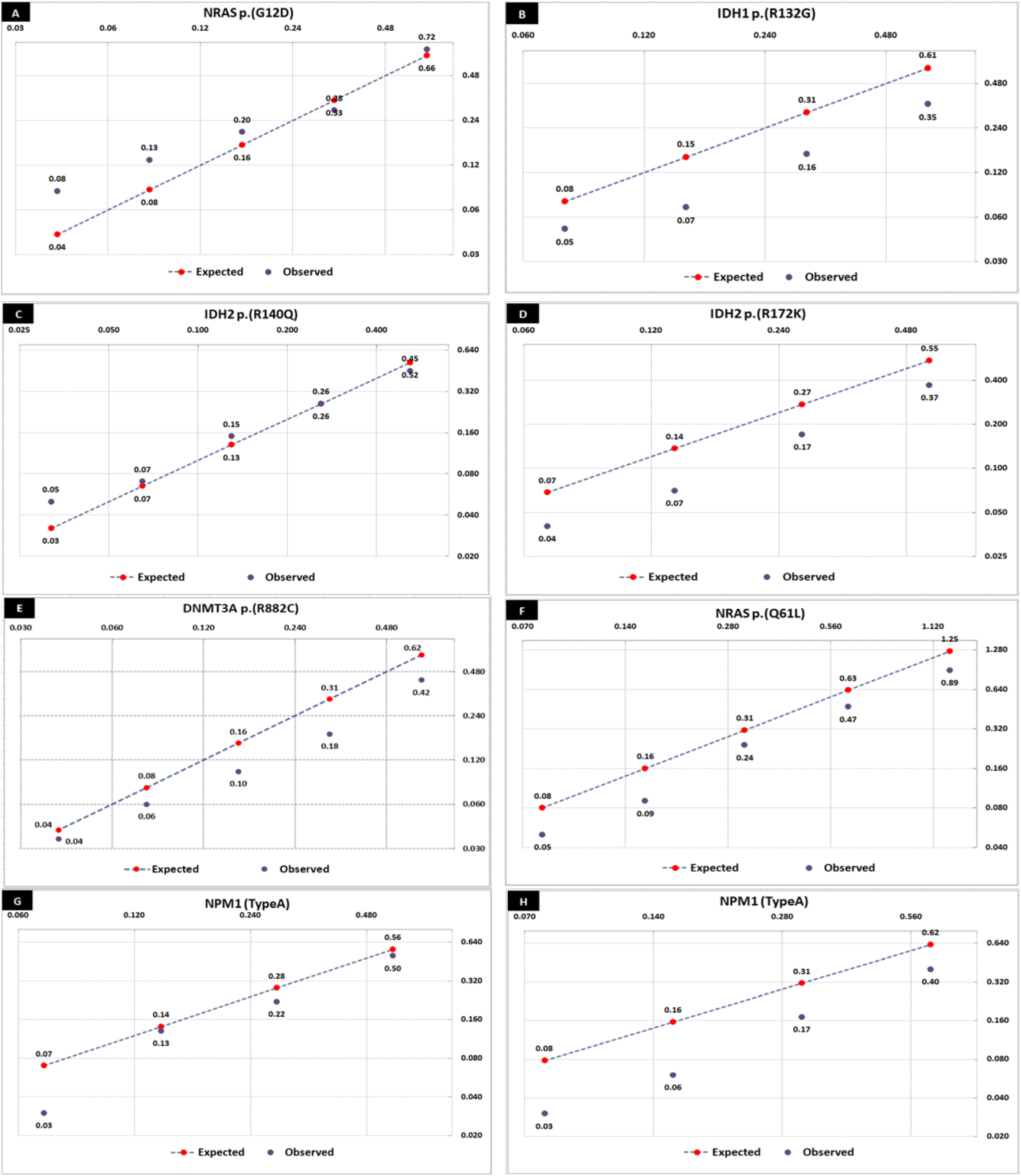
Serial dilutions of OCIAML3 and AML DNA in normal bone marrow. The expected VAF for a mutation was calculated from the original VAF found in undiluted samples.

##### d. Error Modelling

A site and mutation specific error model was setup to ascertain the occurrence of variations observed in the smMIP MRD panel. We sequenced a NA12878 and four normal bone marrow controls to measure sequencing errors using smMIPS as described by Waalkes and colleagues.^2^ As mentioned by Waalkes we discovered a reduction in error rates using UMI based sequencing over standard NGS based sequencing (data not shown). As described, we fitted a β distribution for each base position and probable base substitution error. We observed a higher frequency of C>T and G>A changes consistent with oxidative DNA damage (Supplementary Figure 4) occurring in template DNA before sequencing. Where no variation was detected a 1:15,000 error rate was presumed and a β distribution was modelled. For each variation observed, a site-specific p-value was annotated using these pre-calculated β distributions. Sites with p>0.005 were excluded as artefacts.

**Supplementary Figure 6:**
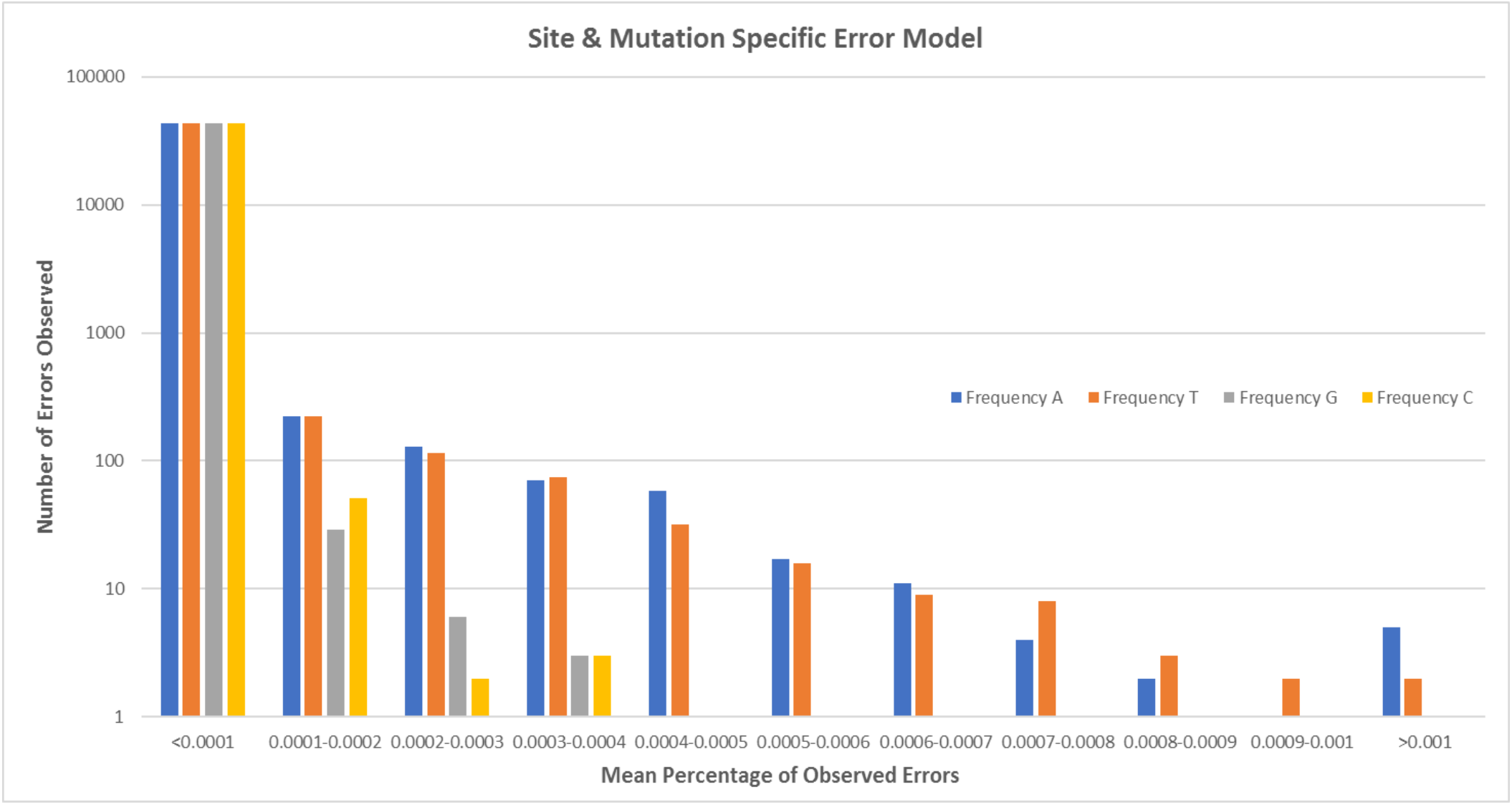
Frequencies of errors observed using a site and mutation specific model.

##### e. Criteria for variant calling using smMIPS MRD assay

i. Variants filtered by focussing on exonic regions (including splicing variants if any) followed by population frequency (<0.01) filtering.
ii. The variant must have been detected at baseline.
iii. Background error modelling at that site must have a *P* value <0.005.
iv. A minimum of 10 alternate variant reads must be present for an SNV.
v. A minimum of 3 alternate variant reads must be present for an indel.
vi. The highest VAF was taken as MRD.
vii. If the highest VAF was in a variant associated with *DNMT3A, TET2* or *ASXL1* (DTA mutation) it was ignored.Supplementary Methods

#### 2. Detection of FLT3-MRD using a one-step PCR assay

##### a. Assay Design

We observed that we could not monitor *FLT3-ITD* using smMIPs. To overcome this issue, we designed a one-step PCR assay that incorporated locus specific primers, dual indices and Illumina adapters in a single step (Supplementary Table 2). The primers were designed to amplify common internal tandem duplication site (chr13: 28608024 – 28608353).^8^ The assay was setup using 600ng of genomic DNA as template. The below primers (50nM each) were added to 12.5¼1 of HotStarTaq Master Mix (Qiagen, Hilden, Germany) to setup a 25 μL reaction. Amplification was carried out under the following conditions; (Denaturation: 95°C-15 minutes; Denaturation: 95°C-60 seconds; Annealing 61°C-60 seconds; Extension 72°C-60 seconds; 35 cycles; final extension 72°C-45 seconds). The final library was size selected using Agencourt AMPure XP beads (Beckman Coulter Inc., California, USA) and sequenced on an Illumina MiSeq (V2, 250PE chemistry). Each sample was allotted 1.1 million reads.

**Supplementary Table 3:**
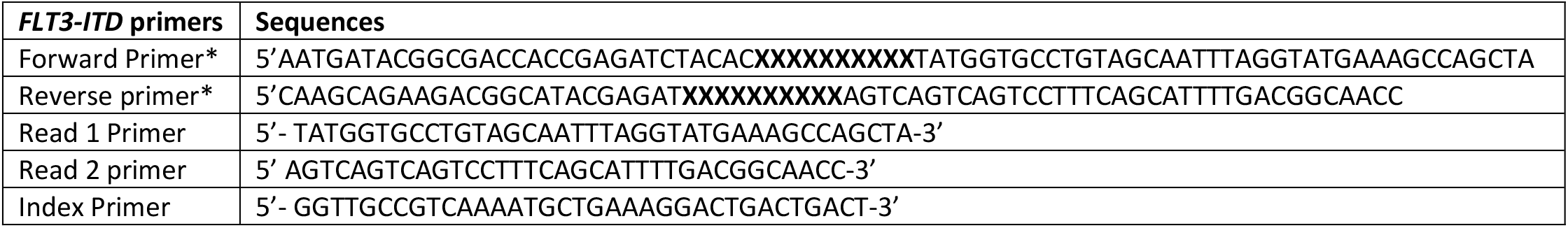
Adapter tagged FLT3-ITD primers *-XXXXXXXXXX represents 10 bp sample specific index sequence.

##### b. Data Analysis and Limit of Detection

We adopted a recently described algorithm^9^ for accurate detection of FLT3-ITD using next generation sequencing. We could demonstrate good correlation (Supplementary Figure 5, inset) between conventional and NGS testing for accurate detection of ITD length in 71 *FLT3-ITD* positive AML. We could validate this assay till a maximum ITD length of 100bp.

##### c. Limit of Detection of FLT3 NGS MRD assay

In order to determine the limit of detection of this assay, we diluted a FLT3-ITD (30bp ITD) positive sample into normal BM. All dilutions were performed in triplicates (Supplementary Figure 5). We could successfully detect this mutation till a lower limit of 0.002% VAF, as shown in Supplementary Figure 5. Based on this data, we established the limit of detection of the FLT3-ITD NGS MRD assay at 0.002. All FLT3-ITD clones >1% VAF were tracked

**Supplementary Figure 7:**
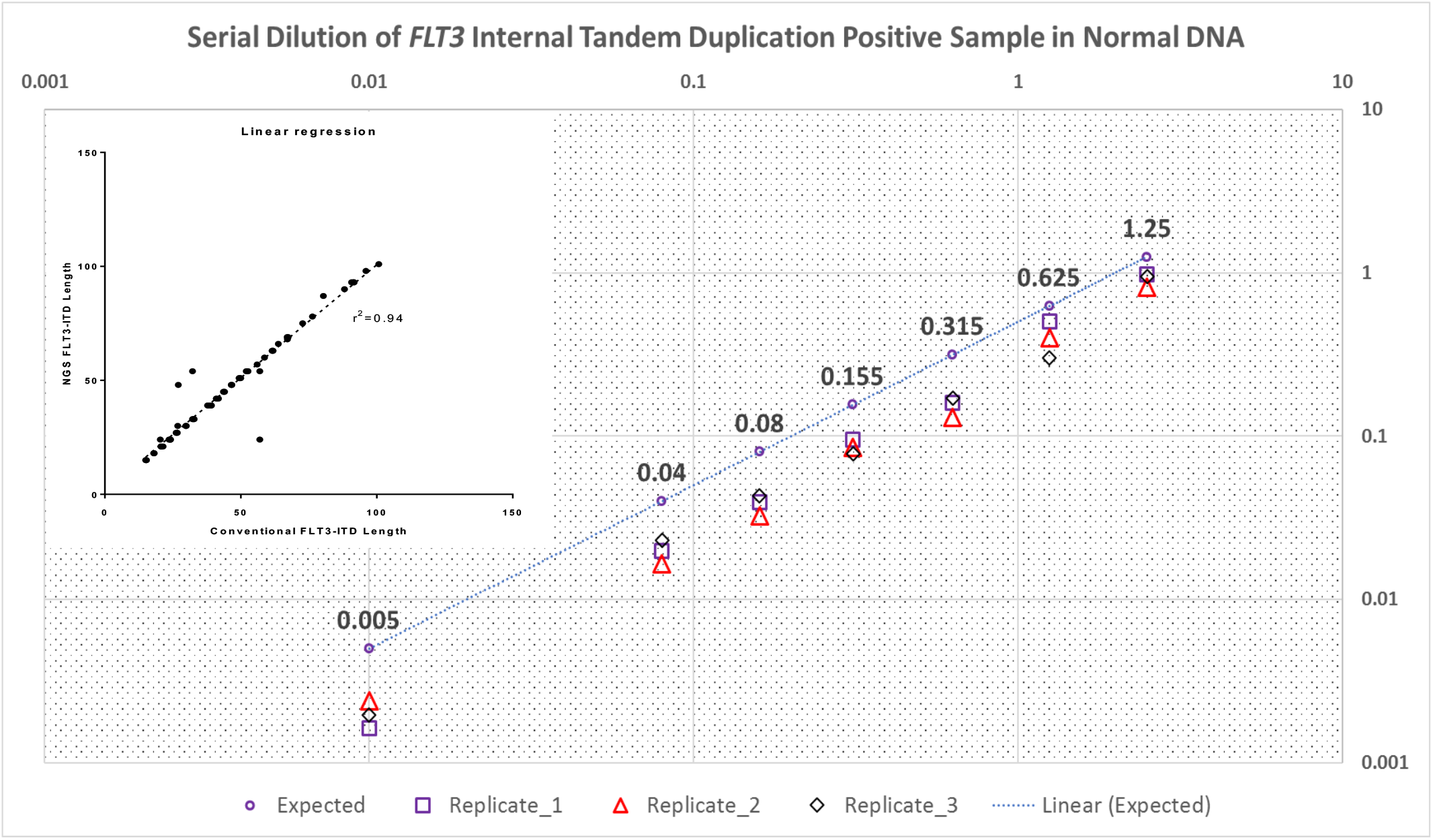
Correlation of *FLT3-ITD* length by capillary (conventional testing) electrophoresis as compared to NGS assay (Inset). Serial dilution of a FLT3-ITD positive sample in normal DNA demonstrates the limit of detection (in triplicates) to be 0.002%.

**Supplementary Table 4:** Distribution of MRD results measured by FCM and NGS into cytogenetic risk categories. FCM: Flow Cytometry; NGS: Next generation sequencing. Chi Squared test did not reveal a statistical significance of MRD results distributed by cytogenetic risk groups.

| Cytogenetic Risk | Post Induction FCM MRD |  | Post Induction NGS MRD |  |
| --- | --- | --- | --- | --- |
|  | MRD Positive | MRD Negative | MRD Positive | MRD Negative |
| **Favorable** | 21 (23.86%) | 27 (24.11%) | 30 (21.58%) | 17 (29.82%) |
| **Intermediate** | 55 (62.50%) | 80 (71.43%) | 99 (71.22%) | 34 (59.65%) |
| **Poor Risk** | 12 (13.64%) | 5 (4.46%) | 10 (7.19%) | 6 (10.53%) |
| **Total** | 88 | 112 | 139 | 57 |

**Supplementary Figure 8:**
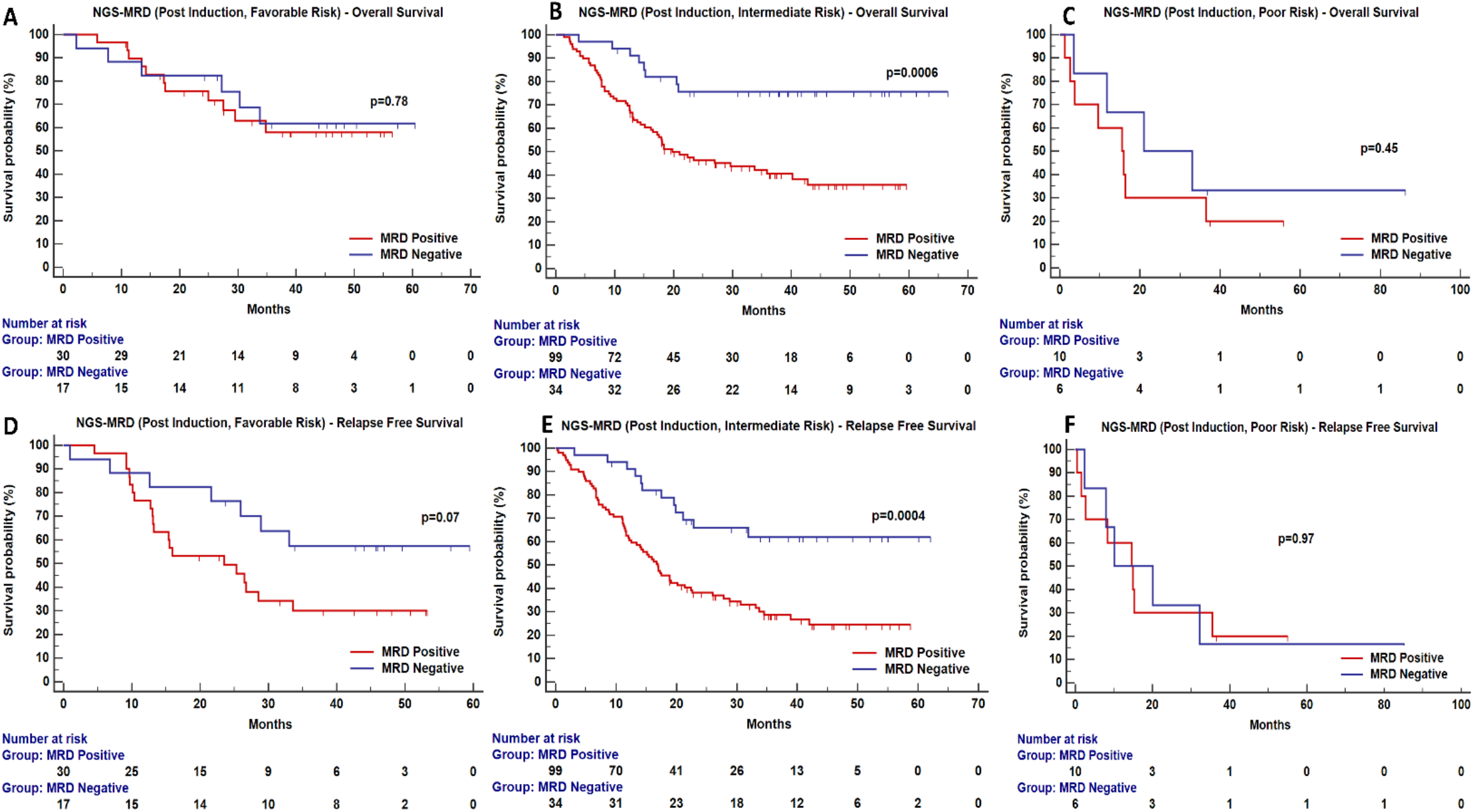
Clinical relevance of NGS-MRD and their influence on overall survival (OS) and relapse free survival (RFS) when patients are grouped by cytogenetic risk categories.

**Supplementary Figure 9:**
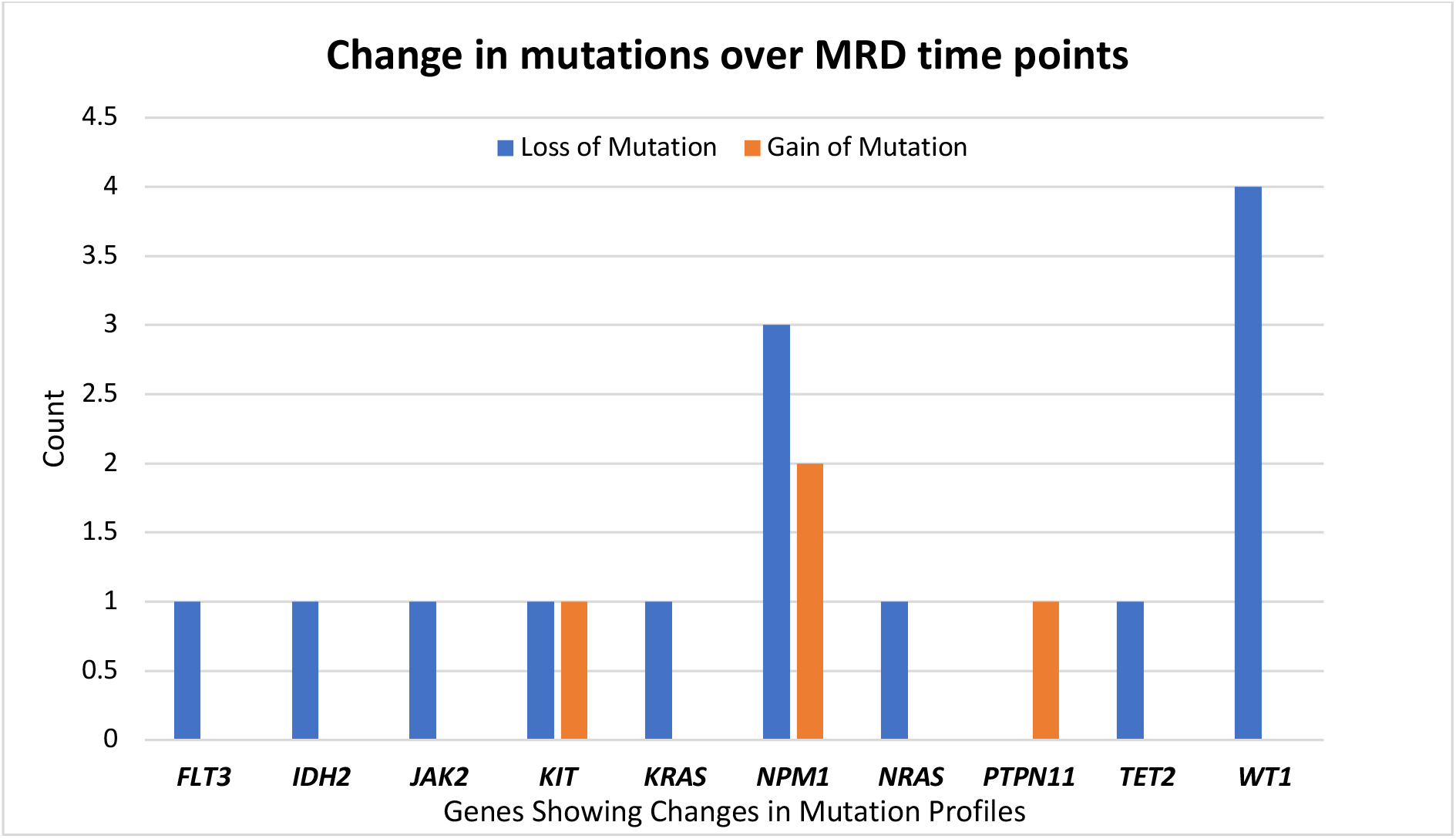
Change in mutations over MRD time points amongst paired PI and PC samples.

**Supplementary Figure 10:**
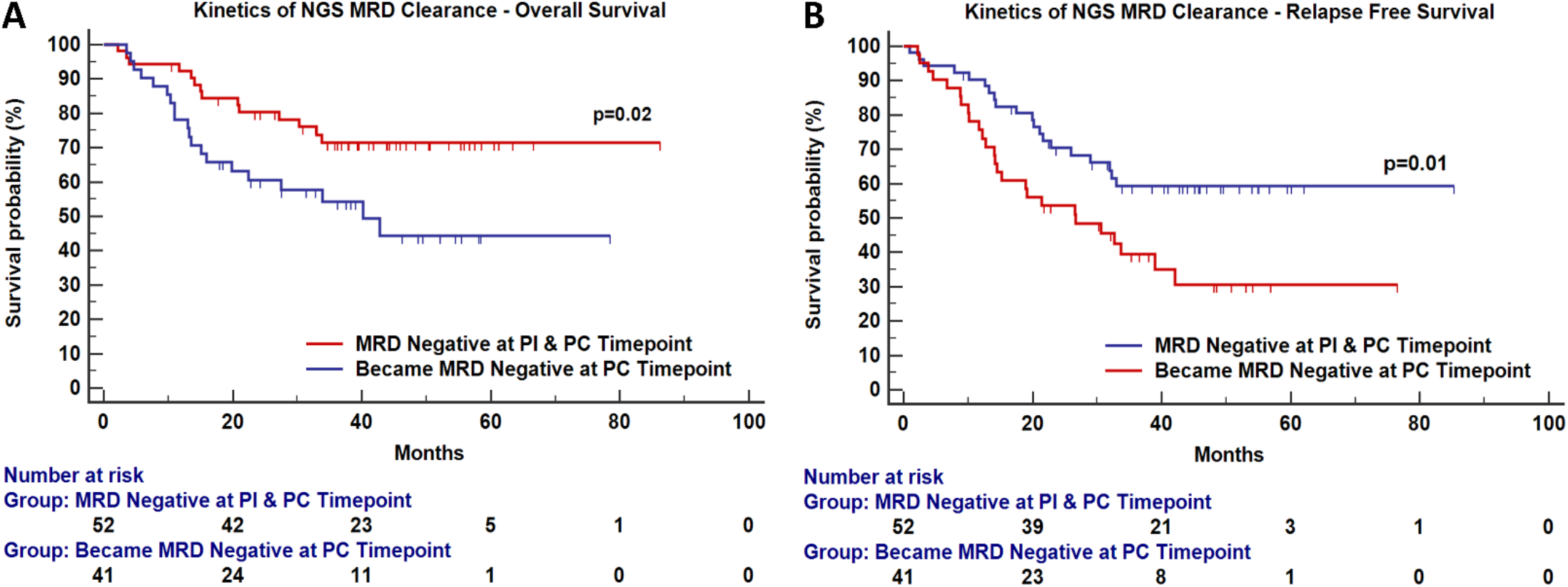
Clinical relevance of kinetics survival (OS) and relapse free survival (RFS).

**Supplementary Figure 11:**
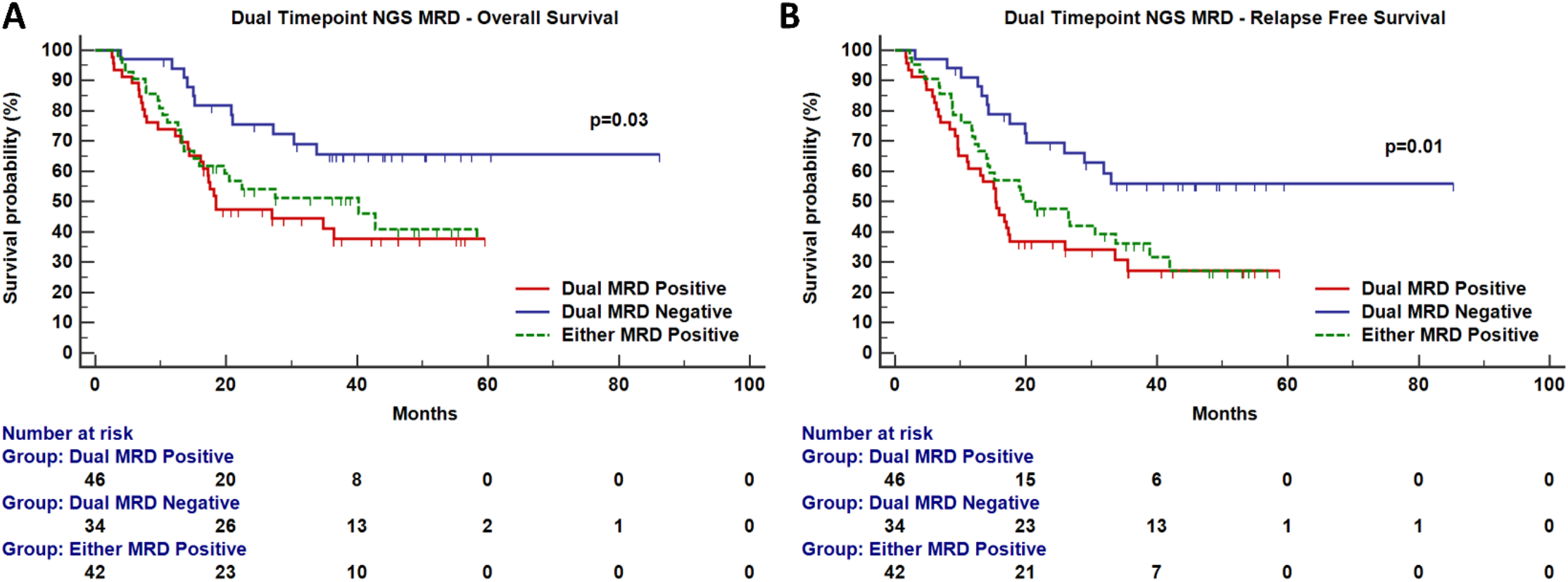
Clinical relevance paired MRD sampling and influence on overall survival (OS) and relapse free survival (RFS). Patients who persistently harboured MRD had a significantly inferior OS [HR- 2.48; 95% CI- 1.33 to 4.61; (p=0.03)] and RFS [HR- 2.55; 95% CI- 1.45 to 4.48; (p=0.01)] as compared to patients who were MRD negative at both time points.50%

**Supplementary Figure 12:**
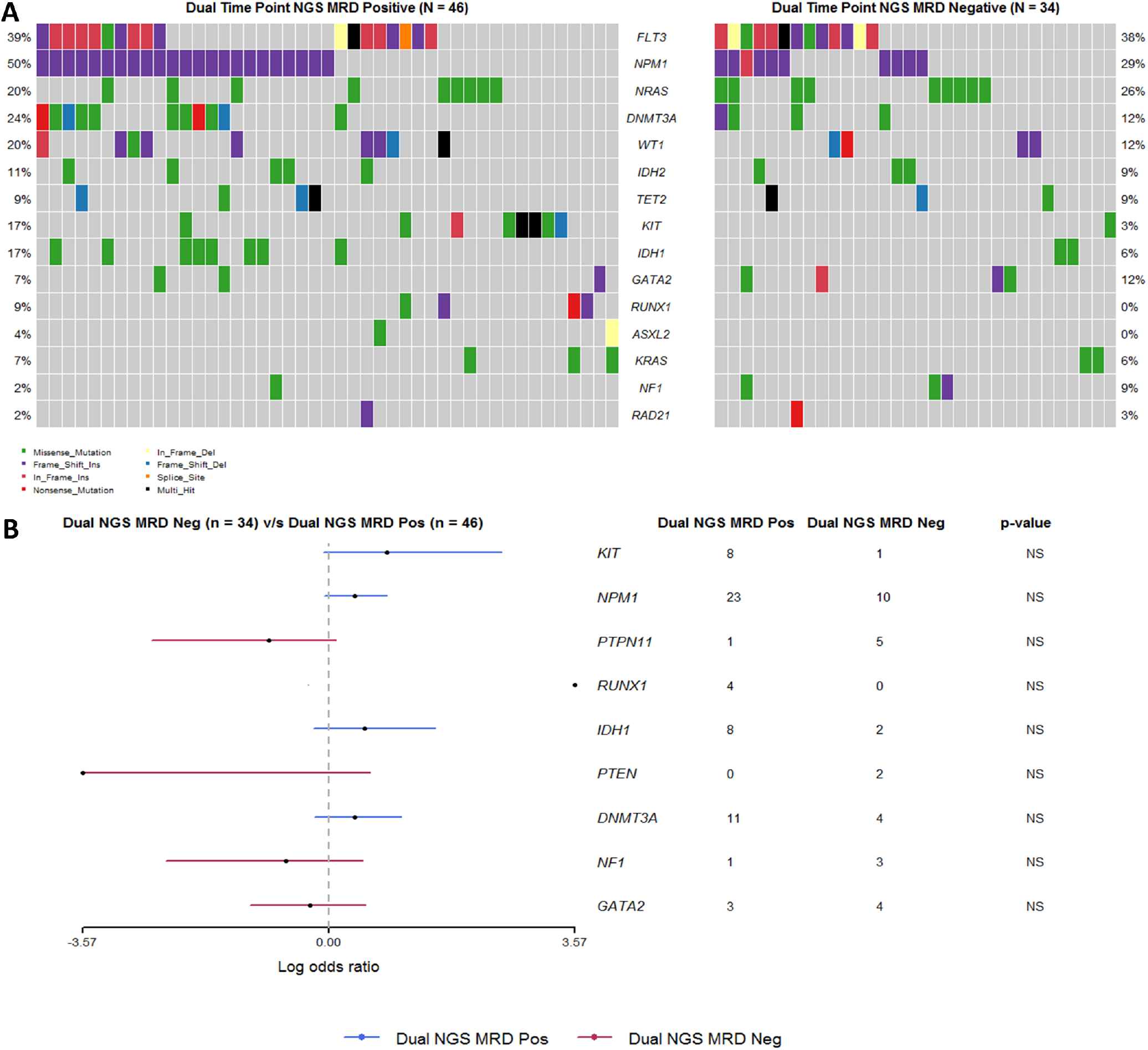
Comparative oncoplot (A) of patients who were NGS-MRD positive at both time points as compared to patients who were persistently MRD negative. Commonly occurring mutations are highlighted here. Forest plot (B) fails to demonstrate a genetic difference between these two groups.

#### 3. Orthogonal MRD detection of NPM1 mutations using NPM1 MRD assay

**Supplementary Figure 13:**
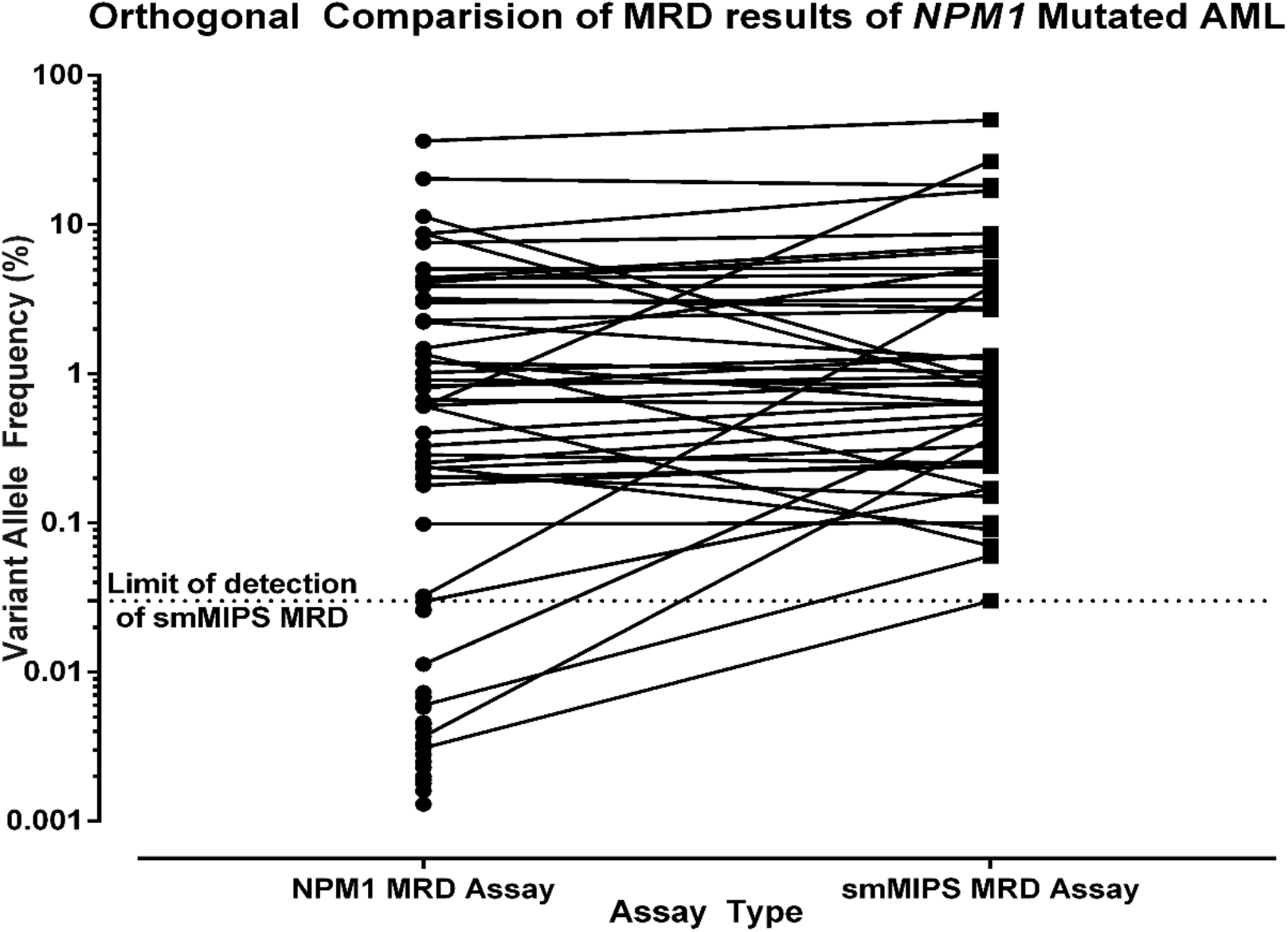
Cross validation of smMIPS MRD cases (n=75; *NPM1* mutated AML) detected by *NPM1* NGS MRD assay. Since the axis is logarithmic, zero values (n=44) have been omitted.

Orthogonal MRD testing was performed in 75 MRD samples (75 out of 323 MRD samples; 23.2%). These were detected using a previously published ultradeep sequencing based MRD assay for *NPM1* mutated AML.^7^ smMIPS MRD positive *NPM1* mutated AML cases (45 out of 75; 60%) could be detected by the *NPM1* NGS MRD assay. For the rest of the cases, *NPM1* mutation was either negative or detected at a threshold below the smMIPS MRD assay limit. For smMIPS MRD positive cases the median MRD value was 0.86% as compared to 0.82% for *NPM1* NGS MRD (at a LOD cut-off of 0.03%).

#### 4. FCM based AML-MRD

**Supplementary Figure 14:**
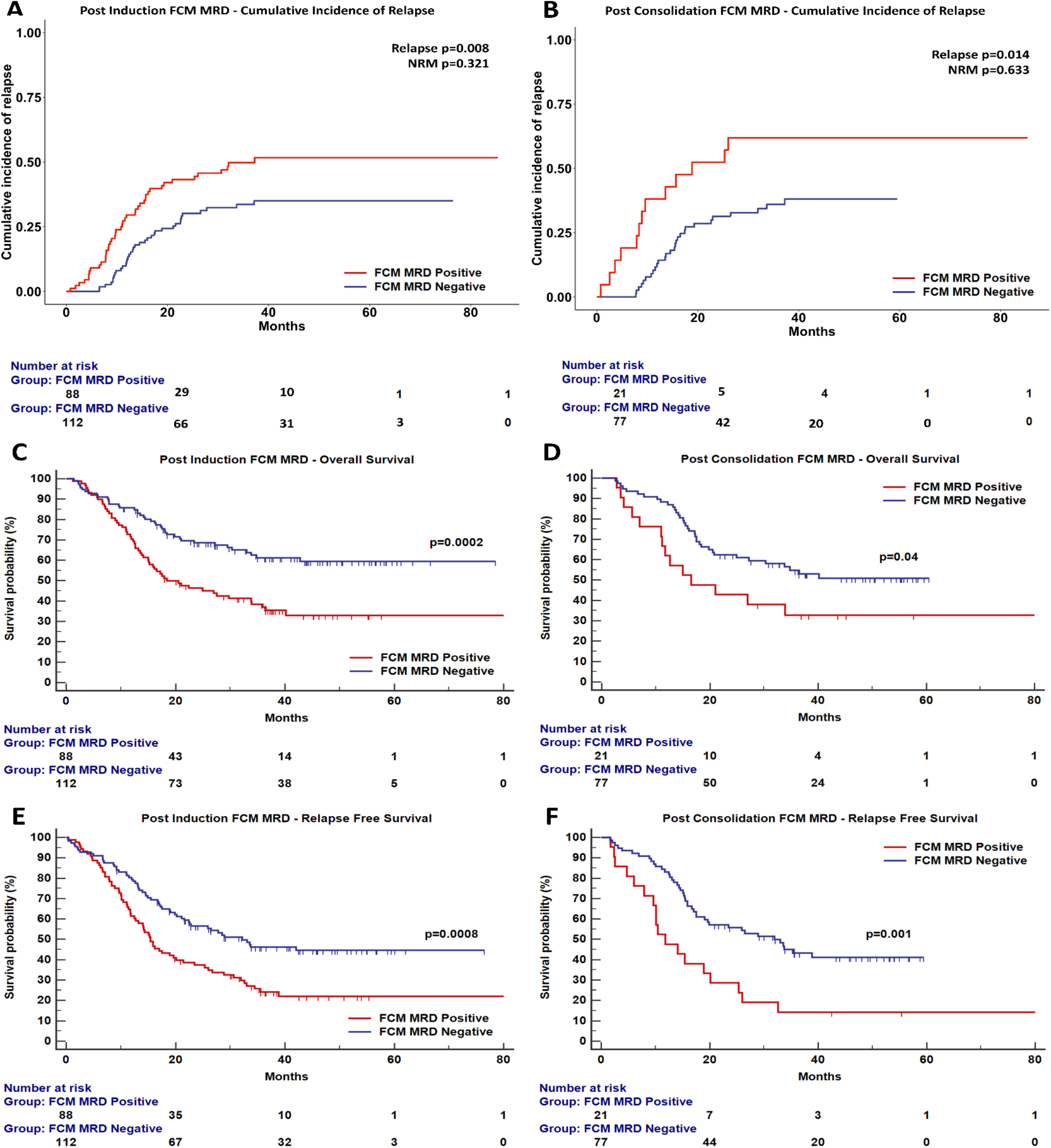
Survival analysis of adult AML patients by flow cytometric MRD assessment (FCM-MRD) after induction and consolidation phases of chemotherapy.

**Supplementary Figure 15:**
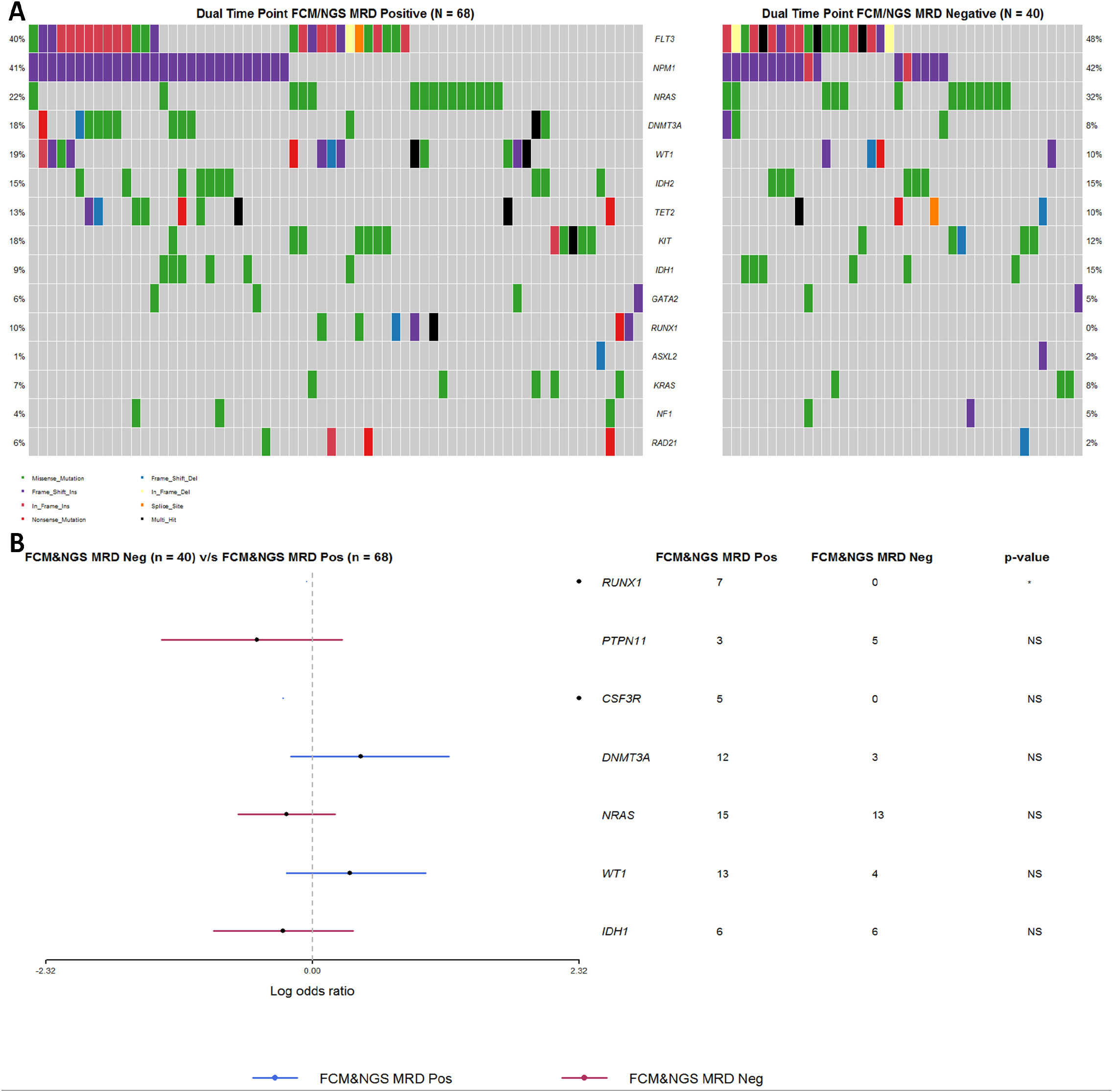
Comparison of genetic profiles between FCM-MRD and NGS-MRD groups (FCM+NGS+ vs FCM-NGS-) at PI timepoint. Comparison is performed using Fischers exact test on all genes between two cohorts *(RUNX1* mutation, p=0.04).

**Supplementary Table 5:** Clinical, cytogenetic risk, molecular risk stratification, NGS MRD coverage, NGS MRD and Flow MRD results and outcome of patients who are FCM MRD positive but NGS MRD negative. NA: MRD not performed

| UPN | Baseline Mutations | Number Covered by NGS Panel | Mutation_Details | FISH | Karyotyping | Cytogenetic Risk | PI NGS-MRD Consensus Coverage | PI NGS-MRD (VAF) | Post Induction NGS MRD | PC NGS-MRD Consensus Coverage | PC NGS-MRD (VAF) | Post Consolidation NGS MRD | Post Induction Flow MRD | Post Consolidation Flow MRD | Survival Status |
| --- | --- | --- | --- | --- | --- | --- | --- | --- | --- | --- | --- | --- | --- | --- | --- |
| Case 1 | 4 | 3 | NRAS:NM_002524:exon2:c.G35A:p.G12D<br>KIT:NM_000222:exon17:c.G2446T:p.D816Y<br>WT1:NM_024426:exon7:c.C1142A:p.S381X<br>FLT3:NM_004119:exon16:c.C2028A:p.N676K | inv(16) and trisomy 8 | Not available | Favorable | 11427 | NRAS- 0.5 | Positive | 18011 |  | Negative | Positive | Positive | Relapsed |
| Case 2 | 1 | 1 | KIT:NM_000222:exon17:c.G2446C:p.D816H | RUNX1-RUNX1T1 fusion: t(8;21) | Not available | Favorable |  | NA | NA | 14314 |  | Negative | Positive | Positive | Relapsed |
| Case 3 | 2 | 1 | NRAS:NM_002524:exon2:c.G38T:p.G13V<br>ATRX:NM_000489:exon9:c.G1948A:p.E650K | RUNX1-RUNX1T1 fusion: t(v;8;21) | Not available | Favorable | 9958 |  | Negative | 17653 |  | Negative | Positive | Negative | Alive |
| Case 4 | 1 | 1 | TET2:NM_001127208:exon6:c.G3799A:p.E1267K | RUNX1-RUNX1T1 fusion: t(8;21) | Not available | Favorable | 20295 |  | Negative | 8716 |  | Negative | Positive | Negative | Alive |
| Case 5 | 5 | 1 | PTEN:NM_000314:exon7:c.697_697delinsGGG<br>DNMT3A:NM_022552:exon4:c.C446T:p.A149V<br>NRAS:NM_002524:exon3:c.A183C:p.Q61H<br>RAD21:NM_006265:exon8:c.C877T:p.Q293X | RUNX1-RUNX1T1 fusion: t(8;21) | 46,XX,<br>t(8;21)(q22;q22)[9]<br>46,XX[2] | Favorable | 7271 |  | Negative | 8627 |  | Negative | Positive | Negative | Died without Relapse |
| Case 6 | 1 | 1 | U2AF1:NM_006758:exon2:c.C101A:p.S34Y | Negative | 46,XY [20] | Intermediate | 16126 | U2AF1- 0.26 | Positive | 9788 |  | Negative | Positive | Positive | Relapsed |
| Case 7 | 1 | 1 | NRAS:NM_002524:exon2:c.G35A:p.G12D | Negative | 46,XY[20] | Intermediate | 16514 |  | Negative |  | NA | NA | Positive | Negative | Alive |
| Case 8 | 1 | 1 | BCOR:NM_001123383:exon4:c.G2503A:p.A835T | Negative | Not available | Intermediate | 20553 |  | Negative | 19283 |  | Negative | Positive | Positive | Relapsed |
| Case 9 | 1 | 1 | FLT3-ITD | Negative | Not available | Intermediate | 1151064 (FLT3-ITD) |  | Negative | 1406146 (FLT3-ITD) |  | Negative | Positive | NA | Alive |
| Case 10 | 2 | 2 | NRAS:NM_002524:exon2:c.G38A:p.G13D<br>FLT3-ITD | MLL translocation | Not available | Intermediate | 15532 |  | Negative | 14215 , (512548 FLT3-ITD) | FLT3 ITD- 0.006 | Positive | Positive | Positive | Died without Relapse |
| Case 11 | 1 | 1 | KRAS:NM_004985:exon4:c.G436A:p.A146T | Negative | Not available | Intermediate | 14179 |  | Negative | 12020 |  | Negative | Positive | NA | Relapsed |
| Case 12 | 3 | 3 | TET2:NM_001127208:exon3:c.778_793AGTGTC<br>NPM1:NM_002520:exon11:c.859_860insTCTG:p.L287fs<br>PTPN11:NM_002834:exon13:c.G1530T:p.Q510H | Negative | Not available | Intermediate | 13335 |  | Negative | 14160 |  | Negative | Positive | Negative | Died without Relapse |
| Case 13 | 1 | 1 | WT1:NM_024426:exon7:c.1109_1109delinsCC | Negative | 46,XY, del(6)(q23.3)[3]<br>46,XY [7] | Intermediate | 12200 |  | Negative | 14645 |  | Negative | Positive | Negative | Relapsed |
| Case 14 | 2 | 2 | NPM1:NM_002520:exon11:c.859_860insTCTG:p.L287fs<br>IDH2:NM_002168:exon4:c.G419A:p.R140Q | Negative | 46,XX, ?inv(10)<br>(p13;q22) [13] 46,XX [7] | Intermediate | 8511 |  | Negative | 18301 | NPM1- 0.42 | Positive | Positive | Negative | Relapsed |
| Case 15 | 1 | 1 | PTPN11:NM_002834:exon3:c.A172T:p.N58Y | Trisomy 8 | 47,XY,+8[13] 46,XY[2] | Intermediate | 14838 |  | Negative | 22840 |  | Negative | Positive | Negative | Died without Relapse |
| Case 16 | 2 | 1 | NRAS:NM_002524:exon2:c.G38A:p.G13D<br>NF1:NM_001042492:exon34:c.A4462G:p.T1488A | MLL translocation: t(6;11)(q28;q23) | 46,XY,<br>t(6;11)(q27;q23)[3]<br>46,XY,t(6;11),<br>t(9;11)(q22;q23)[4]<br>46,XY[3] | Poor | 8652 |  | Negative | 10573 |  | Negative | Positive | Positive | Relapsed |
| Case 17 | 3 | 3 | IDH1:NM_005896:exon4:c.C394T:p.R132C<br>TP53:NM_001126114:exon5:c.G524A:p.R175H<br>U2AF1:NM_006758:exon6:c.G471C:p.Q157H | Negative | 46,XX,del(X)(p11)[4],<br>del(11)(q23)[6],<br>del(12)(p13)[4],<br>del(18)(p11)[2][cp9]<br>46,XX[3] | Poor | 7925 |  | Negative | 14034 |  | Negative | Positive | Positive | Relapsed |
| Case 18 | 3 | 3 | SF3B1:NM_012433:exon15:c.A2098G:p.K700E<br>GATA2:NM_032638:exon4:c.A890G:p.N297S<br>KRAS:NM_004985:exon2:c.G35A:p.G12D | Monosomy 7 | Not available | Poor | 7281 |  | Negative |  | NA | NA | Positive | Positive | Died without Relapse |
| Case 19 | 1 | 1 | FLT3-ITD | Trisomy 5, 8 and 21 | Not available | Poor | 1469447 (FLT3-ITD) |  | Negative | 1322385 (FLT3-ITD) |  | Negative | Positive | Positive | Relapsed |
| Case 20 | 1 | 1 | GATA2:NM_032638:exon4:c.G952A:p.A318T | Negative | 46,XX,del(5)(q33q35)[3]<br>46,XX, random abnormalities[2]<br>46,XX,[12] | Poor | 1394 |  | Negative | 3917 |  | Negative | Positive | Positive | Alive |

**Supplementary Table 6:**
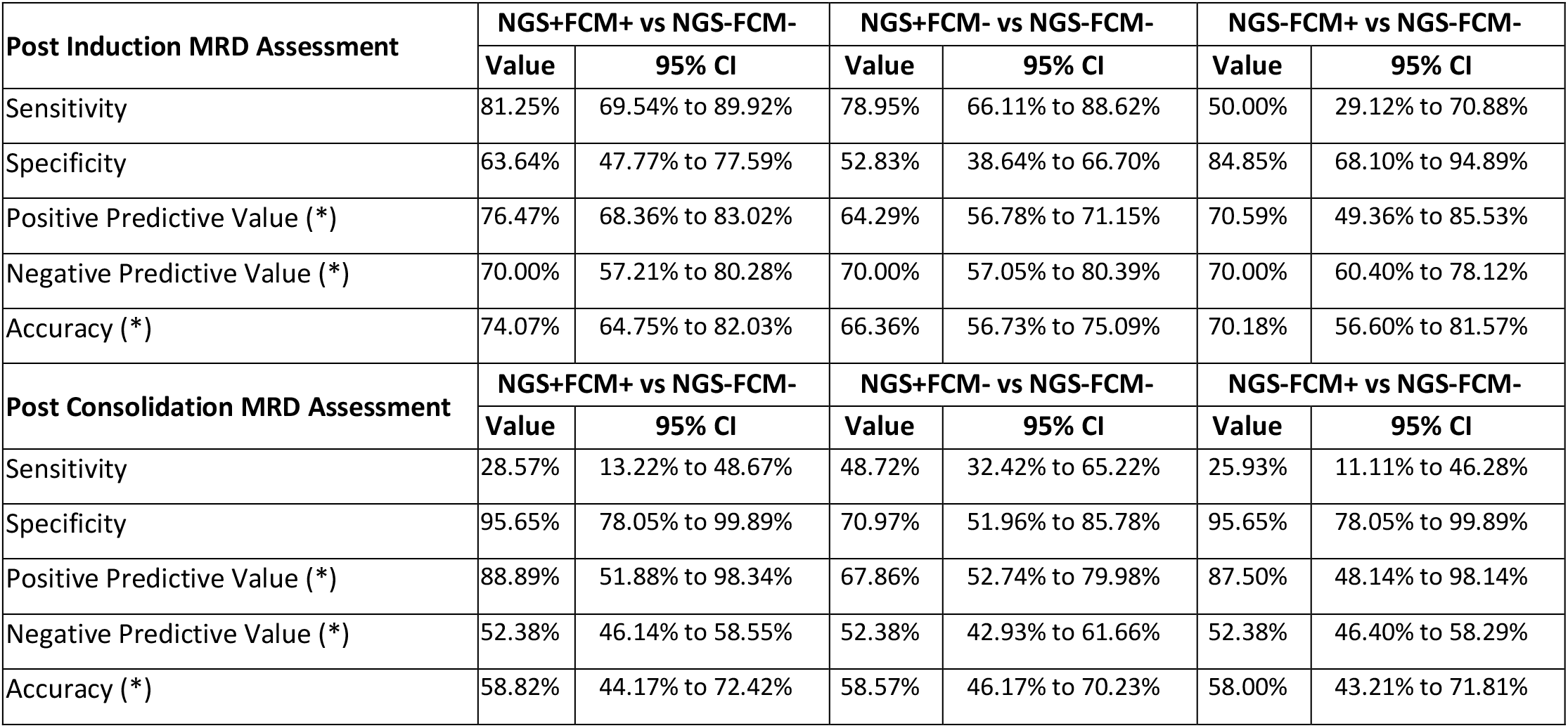
Sensitivity, Specificity, PPV, NPV and Accuracy of both FCM-MRD and NGS-MRD assays.

| Post Induction MRD Assessment | NGS+FCM+ vs NGS-FCM- |  | NGS+FCM- vs NGS-FCM- |  | NGS-FCM+ vs NGS-FCM- |  |
| --- | --- | --- | --- | --- | --- | --- |
|  | Value | 95% CI | Value | 95% CI | Value | 95% CI |
| Sensitivity | 81.25% | 69.54% to 89.92% | 78.95% | 66.11% to 88.62% | 50.00% | 29.12% to 70.88% |
| Specificity | 63.64% | 47.77% to 77.59% | 52.83% | 38.64% to 66.70% | 84.85% | 68.10% to 94.89% |
| Positive Predictive Value (*) | 76.47% | 68.36% to 83.02% | 64.29% | 56.78% to 71.15% | 70.59% | 49.36% to 85.53% |
| Negative Predictive Value (*) | 70.00% | 57.21% to 80.28% | 70.00% | 57.05% to 80.39% | 70.00% | 60.40% to 78.12% |
| Accuracy (*) | 74.07% | 64.75% to 82.03% | 66.36% | 56.73% to 75.09% | 70.18% | 56.60% to 81.57% |
| Post Consolidation MRD Assessment | NGS+FCM+ vs NGS-FCM- |  | NGS+FCM- vs NGS-FCM- |  | NGS-FCM+ vs NGS-FCM- |  |
|  | Value | 95% CI | Value | 95% CI | Value | 95% CI |
| Sensitivity | 28.57% | 13.22% to 48.67% | 48.72% | 32.42% to 65.22% | 25.93% | 11.11% to 46.28% |
| Specificity | 95.65% | 78.05% to 99.89% | 70.97% | 51.96% to 85.78% | 95.65% | 78.05% to 99.89% |
| Positive Predictive Value (*) | 88.89% | 51.88% to 98.34% | 67.86% | 52.74% to 79.98% | 87.50% | 48.14% to 98.14% |
| Negative Predictive Value (*) | 52.38% | 46.14% to 58.55% | 52.38% | 42.93% to 61.66% | 52.38% | 46.40% to 58.29% |
| Accuracy (*) | 58.82% | 44.17% to 72.42% | 58.57% | 46.17% to 70.23% | 58.00% | 43.21% to 71.81% |

**Supplementary Figure 16:**
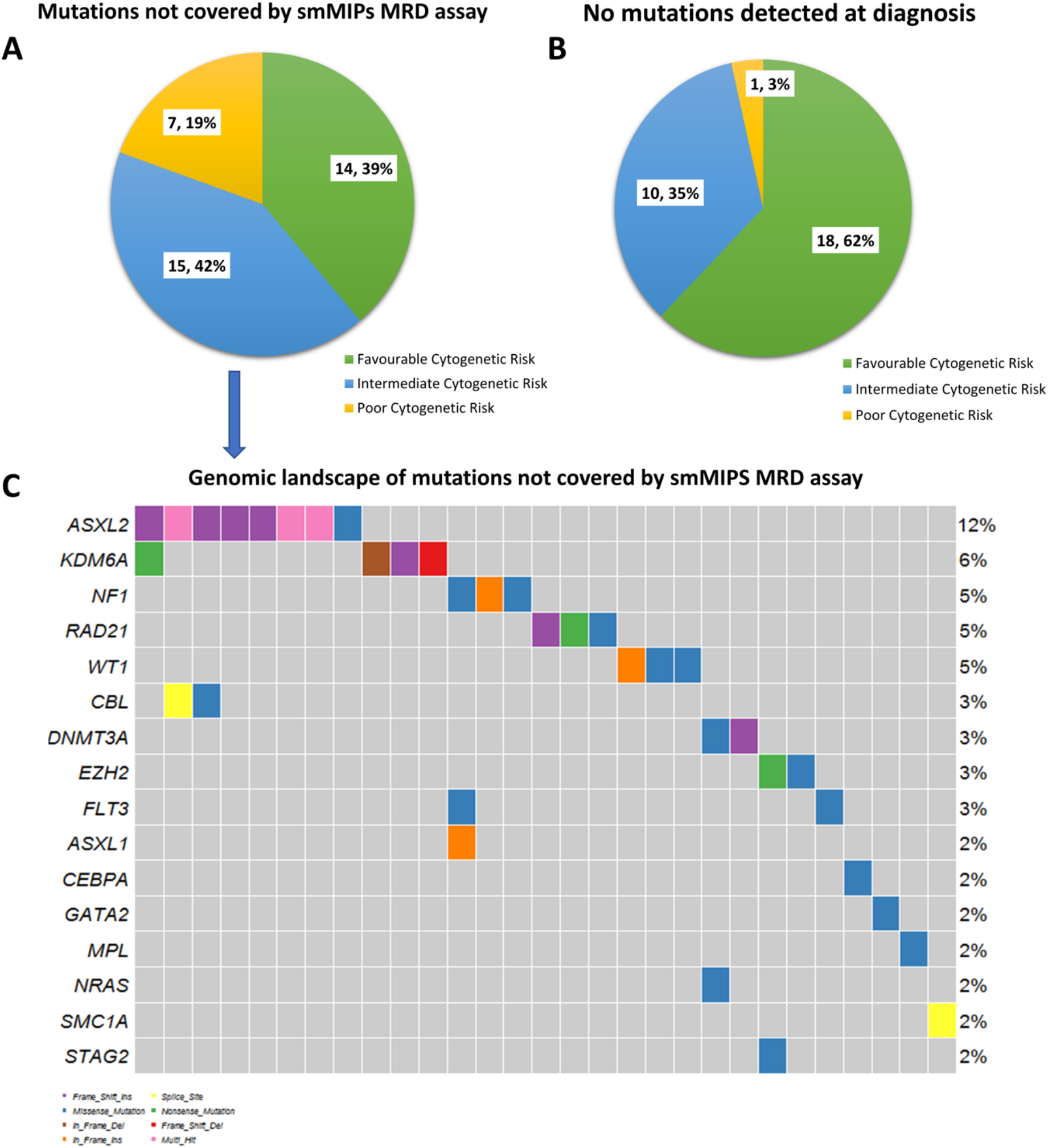
Cytogenetic and genomic landscape of cases which on which NGS-MRD could not be performed. Out of 65 cases, 36 had detectable mutations (A) whereas, no mutations were detected in 29 cases (B). Genomic landscape of cases in which mutations were detected can be seen in a waterfall plot (C). Number of cases are represented in box with percentages in parenthesis. The solitary *CEBPA* mutation seen here was a case sequenced by Illumina Trusight Myeloid Sequencing Panel.

**Supplementary Figure 17:**
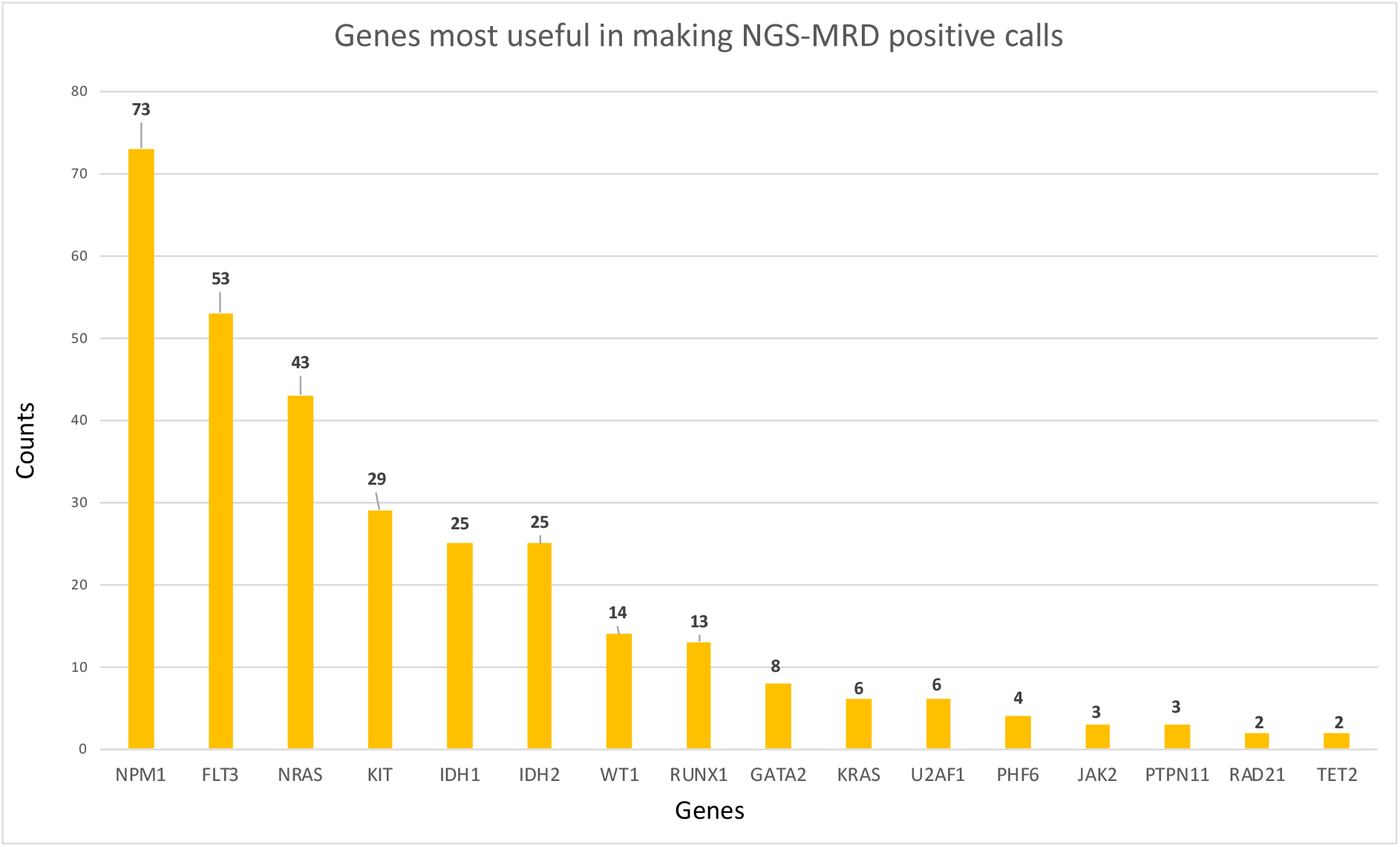
This figure highlights genes that were informative in making a NGS-MRD positive call.

**Supplementary Figure 18:**
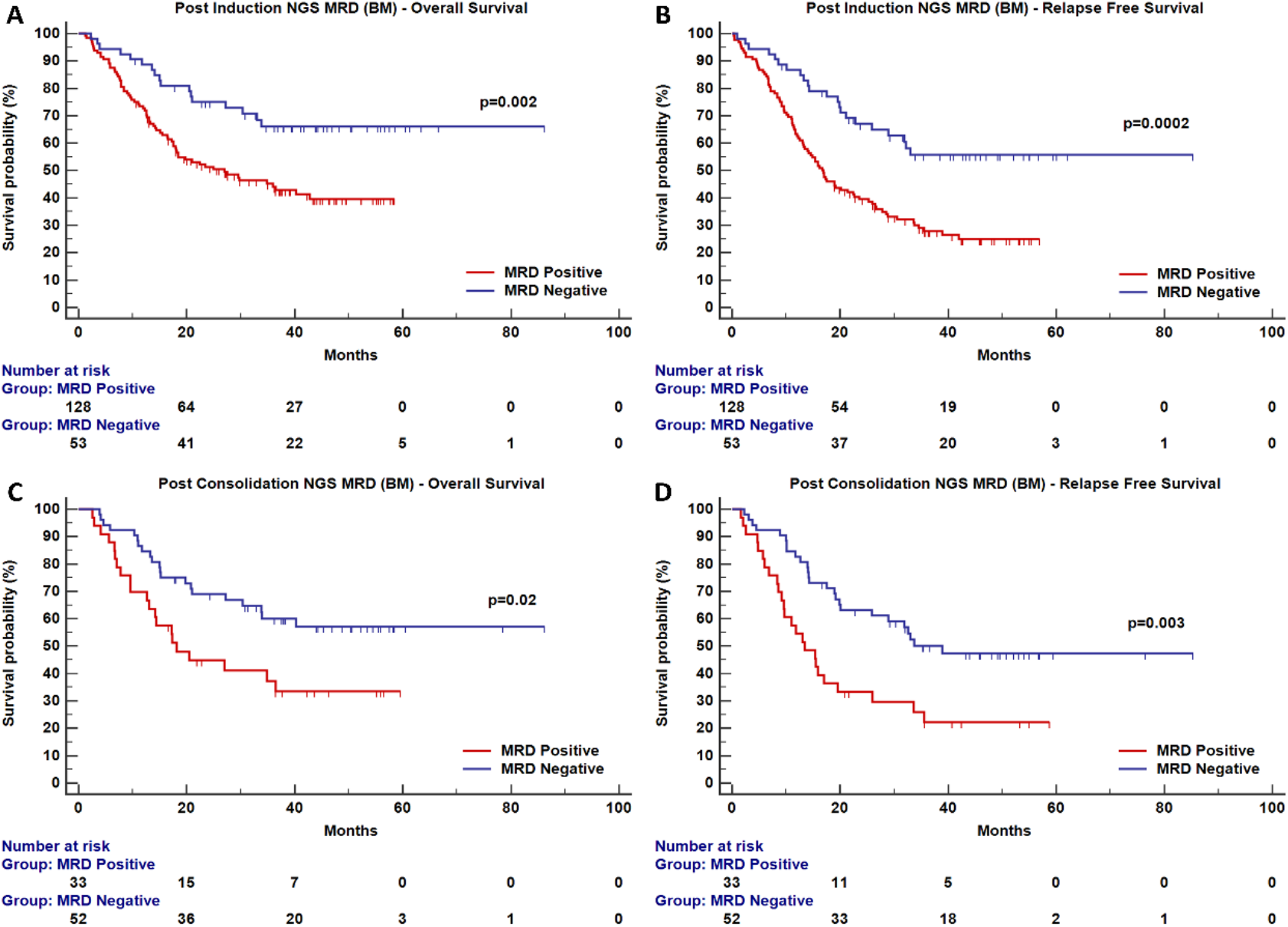
Survival analysis for end of induction and post consolidation using NGS-MRD (After excluding samples sourced from blood).

**Supplementary Figure 19:**
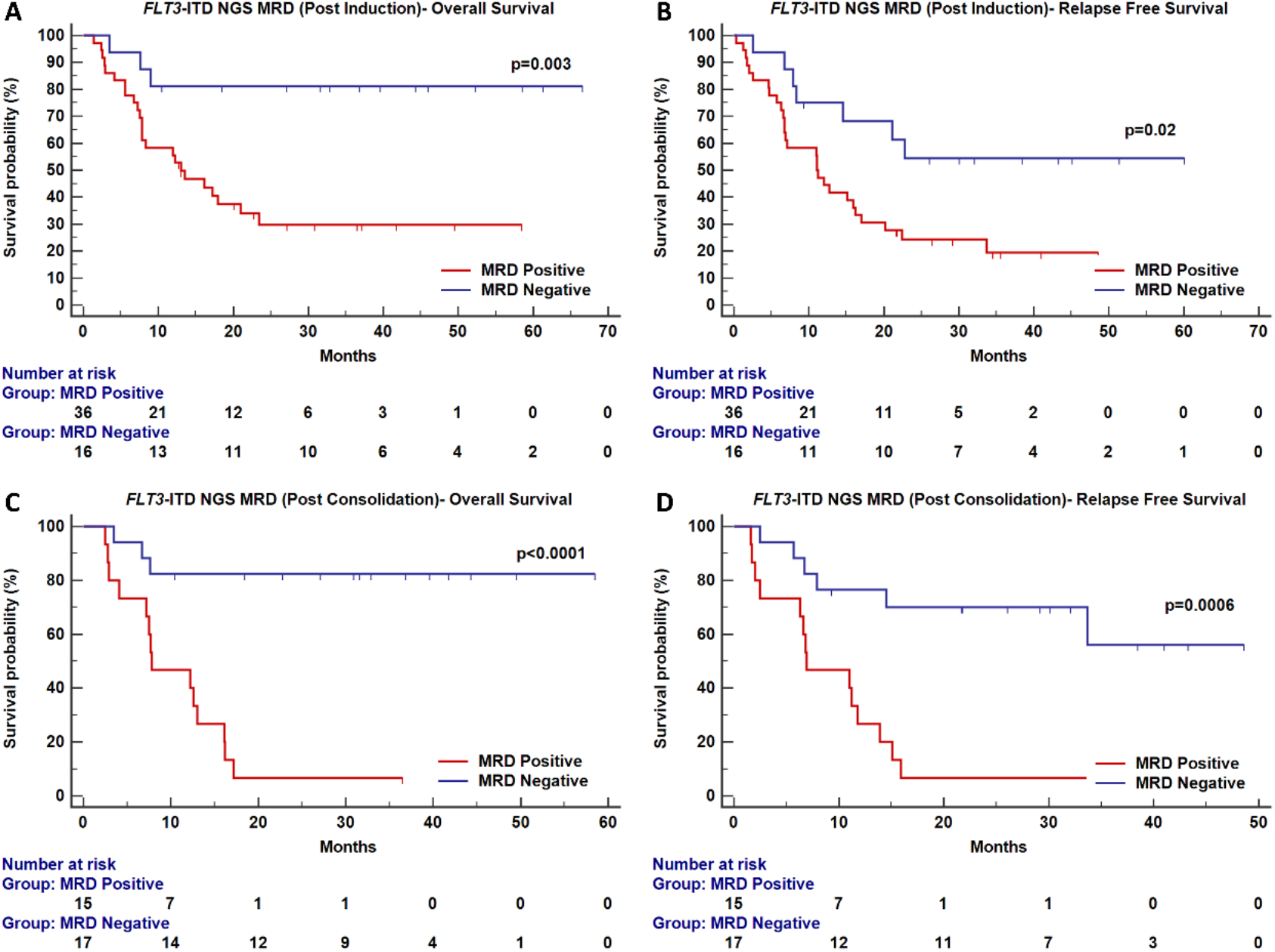
Impact of presence of *FLT3* NGS MRD on overall and relapse free survival at PI and PC.

